# Evaluating symptoms and sensors for menstrual phase estimation beyond bleeding and cycle position

**DOI:** 10.64898/2026.03.31.26349766

**Authors:** Bernhard Specht, Samaher Garbaya, Djamel Khadraoui, Mohammad EL-Khozondar, Reinhard Schneider, Zied Tayeb

**Affiliations:** MyelinZ, Manchester, UK / San Francisco, USA / Leuven, Belgium / Luxembourg; University of Luxembourg, Esch-sur-Alzette, Luxembourg; Luxembourg Centre for Systems Biomedicine (LCSB), Esch-sur-Alzette, Luxembourg; Luxembourg Institute of Science and Technology (LIST), Esch-sur-Alzette, Luxembourg

## Abstract

Whether symptoms and sensor measurements improve menstrual phase estimation beyond bleeding and cycle position remains unclear. We compared input combinations for four-phase estimation in 42 mcPHASES participants using nested feature selection and leave-one-participant-out evaluation against reference phases derived from hormones and menstrual flow. Across two tree-based classifiers, additional symptoms and sensors produced little consistent improvement over bleeding and bleeding-derived cycle position. With a gradient-boosted tree classifier, adding sensors to bleeding, position and symptoms changed macro F1 by +0.17 percentage points (95% confidence interval [-0.61, +1.04]). With sensors alone, features representing recent changes, trends and deviations from participants’ prior averages increased macro F1 from 30.08% to 43.55%. The sensor-only estimates used temporal filtering to incorporate expected phase order and duration using only current and past observations. Fertility had the lowest phase-specific F1, limiting interpretation for fertility-related decisions. Sensor measurements thus provided information about menstrual phase but offered little consistent additional benefit when bleeding and cycle position were already known. Studies evaluating additional symptoms or sensor measurements should include bleeding and bleeding-derived cycle position as baselines to establish their incremental predictive value.

## Introduction

Menstrual phase tracking can draw on bleeding reports, symptom questionnaires and physiological measurements from wearable or continuous monitoring devices. These inputs differ in what they measure and in the effort required to collect them [1]. Understanding which measurements improve phase estimation can guide the design of tracking tools, help reduce unnecessary reporting or sensing, and identify priorities for future data collection. A useful assessment must therefore consider both the information a modality provides on its own and what it adds when simpler observations are already available.

Previous research has linked self-reported symptoms to cycle-length variability [2] and examined glucose and wrist-temperature variation across the cycle or in relation to hormones [3, 4]. Modelling studies have used self-tracked records to label reproductive states [5], wearable measurements to identify menstrual phases [6], and physiological signals to detect the fertile window [7]. A systematic review has also assessed wearable approaches to fertility-window detection [8]. Together, these studies establish a basis for investigating symptoms and sensors in menstrual tracking. However, associations with cycle physiology and successful prediction from combined inputs do not establish how much each measurement improves prediction beyond bleeding and cycle position.

Recent analyses of mcPHASES address related aspects of this question. Nsugbe and Lin compared combinations of hormonal, metabolic and wearable inputs, with normalized cycle day included in the hormone group [9]. Xu et al. reported that wearable-based ovulation inference improved subsequent next-menses timing over static calendar and history references, with gains varying across cycle-irregularity profiles [10]. FemWear investigates wearable representation learning across menstrual and other health tasks [11], while an unreviewed public analysis reports strong phase-classification performance using bleeding-derived cycle timing alone [12]. These findings motivate a more specific measurement-selection question: which non-hormonal input families and sensor categories improve daily four-phase estimation when bleeding and cycle position are already known? Answering this question requires distinguishing cycle timing from non-flow symptoms and comparing inputs within the same cohort and prediction task.

The mcPHASES dataset provides an opportunity for these comparisons by combining self-reports and non-hormonal measurements with reference phases derived from hormone estimates and menstrual-flow boundaries [13, 14]. The source study also examined participants’ experiences of multimodal tracking [1]. Its paired measurements allow input combinations to be evaluated on the same participants and days. Because menstrual flow contributes to the reference definitions, the predictive value of bleeding must be interpreted in that context.

Here, we evaluate the standalone and incremental predictive value of non-hormonal inputs for daily menstrual phase estimation. We ask which input families and sensor categories add value beyond bleeding and position, how the representation of changes over time affects prediction when bleeding and position are unavailable, and how findings depend on report availability and temporal filtering. By identifying where additional measurements help and where their benefit remains uncertain, this analysis aims to inform measurement selection and priorities for further investigation.

## Results

### Incremental value of symptoms and sensors

Adding symptoms and sensors to bleeding and cycle position produced small mean changes in macro F1, with confidence intervals spanning zero for both classifieronly pipelines. With history features, CatBoost classifier macro F1 was 62.86% for bleeding and position and 63.28% for all four input families; the corresponding Extra Trees scores were 64.23% and 64.20% (Table 1). Accuracy and performance across all input-family combinations are reported in Table S1 and Figure S2.

**Table 1:** Phase-classification performance by input combination. Values are participant-average macro F1 (%) with nominal 95% participant-bootstrap confidence intervals in parentheses. 3 seeds are averaged within each of 42 participants; all models use history encoding. Bold indicates the highest displayed mean within each model and prediction method, including ties; it does not imply a statistically significant difference. All four input families comprise bleeding, position, symptoms and sensors.

| Inputs | Classifier only | Classifier + HSMM |
|---|---|---|
| **CatBoost** |  |  |
| Cycle position | 54.8 (48.6, 60.6) | 53.9 (47.9, 59.9) |
| Bleeding + position | 62.9 (58.4, 67.1) | 64.0 (59.7, 68.1) |
| Bleeding + position + symptoms | 63.1 (58.7, 67.3) | 63.4 (58.9, 67.7) |
| All four input families | **63.3** (59.0, 67.3) | **64.5** (60.2, 68.6) |
| **Extra Trees** |  |  |
| Cycle position | 53.7 (47.5, 59.7) | 54.5 (48.2, 60.5) |
| Bleeding + position | **64.2** (60.3, 68.0) | 63.9 (59.7, 67.7) |
| Bleeding + position + symptoms | **64.2** (60.2, 68.0) | **64.0** (59.8, 68.1) |
| All four input families | **64.2** (60.2, 68.0) | 63.9 (59.8, 67.8) |

Adding symptoms to bleeding and position changed CatBoost classifier macro F1 by +0.24 percentage points (95% interval [-0.44, +0.96]). Adding sensors to bleeding, position and symptoms changed it by +0.17 points ([-0.61, +1.04]). The corresponding Extra Trees changes were -0.07 ([-1.05, +0.90]) and +0.04 points ([-0.92, +0.95]). With hidden semi-Markov model (HSMM) filtering, adding sensors to bleeding, position and symptoms improved CatBoost macro F1 by +1.09 points (nominal 95% interval [+0.39, +1.83]), but the difference did not remain significant after Holm adjustment (*p* = 1.00). Extra Trees did not show the same improvement (Figure 1).

**Figure 1:**
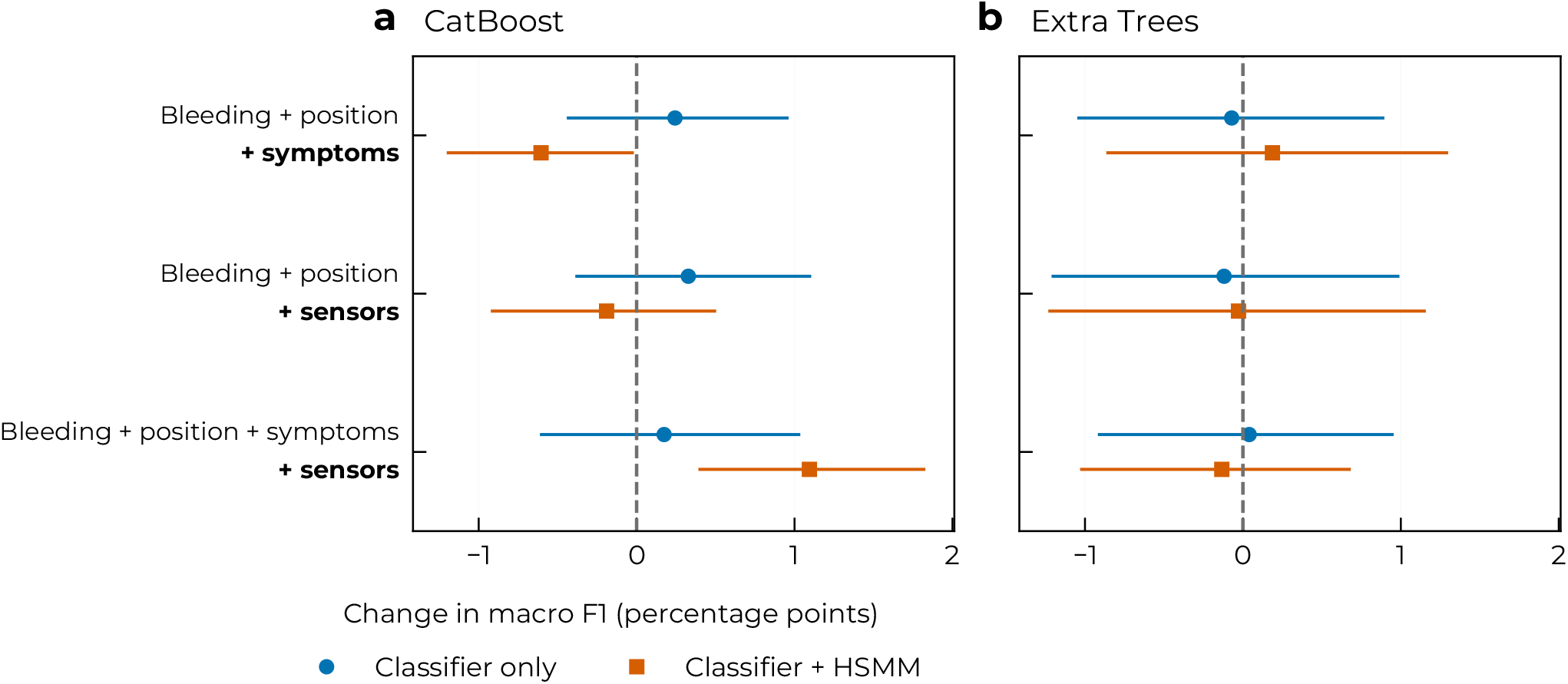
Incremental value of symptoms and sensors. **a**,**b**, Change in macro F1 after retraining CatBoost (a) and Extra Trees (b) with the added inputs. Baselines are specified by row; position derives from bleeding onset. Circles denote classifier-only predictions and squares classifier predictions with forward HSMM filtering. All models use history features. Points show mean paired changes across 42 held-out participants, averaging 3 seeds within participant. Bars show 95% paired participant-bootstrap intervals (5,000 resamples), without multiplicity adjustment. Positive values favour added inputs; dashed lines indicate no change.

The matched comparison with the pipeline without feature selection showed a similar overall pattern for classifier-only predictions. With nested feature selection, adding symptoms and sensors together to bleeding and position changed macro F1 by +0.42 percentage points (95% interval [-0.55, +1.51]) for CatBoost and -0.03 points ([-1.15, +1.03]) for Extra Trees (Figure S8).

Individual sensor categories produced positive and negative mean changes depending on the baseline. Adding resting heart rate to bleeding and position increased classifier-only macro F1, with an unadjusted 95% confidence interval excluding zero. Temperature and heart rate added to bleeding, position and symptoms showed nominal improvements with HSMM filtering. None of these additions remained significant after Holm adjustment (Figure 2). Phase-specific additions showed further variation across the four phases (Figures S4 and S5).

**Figure 2:**
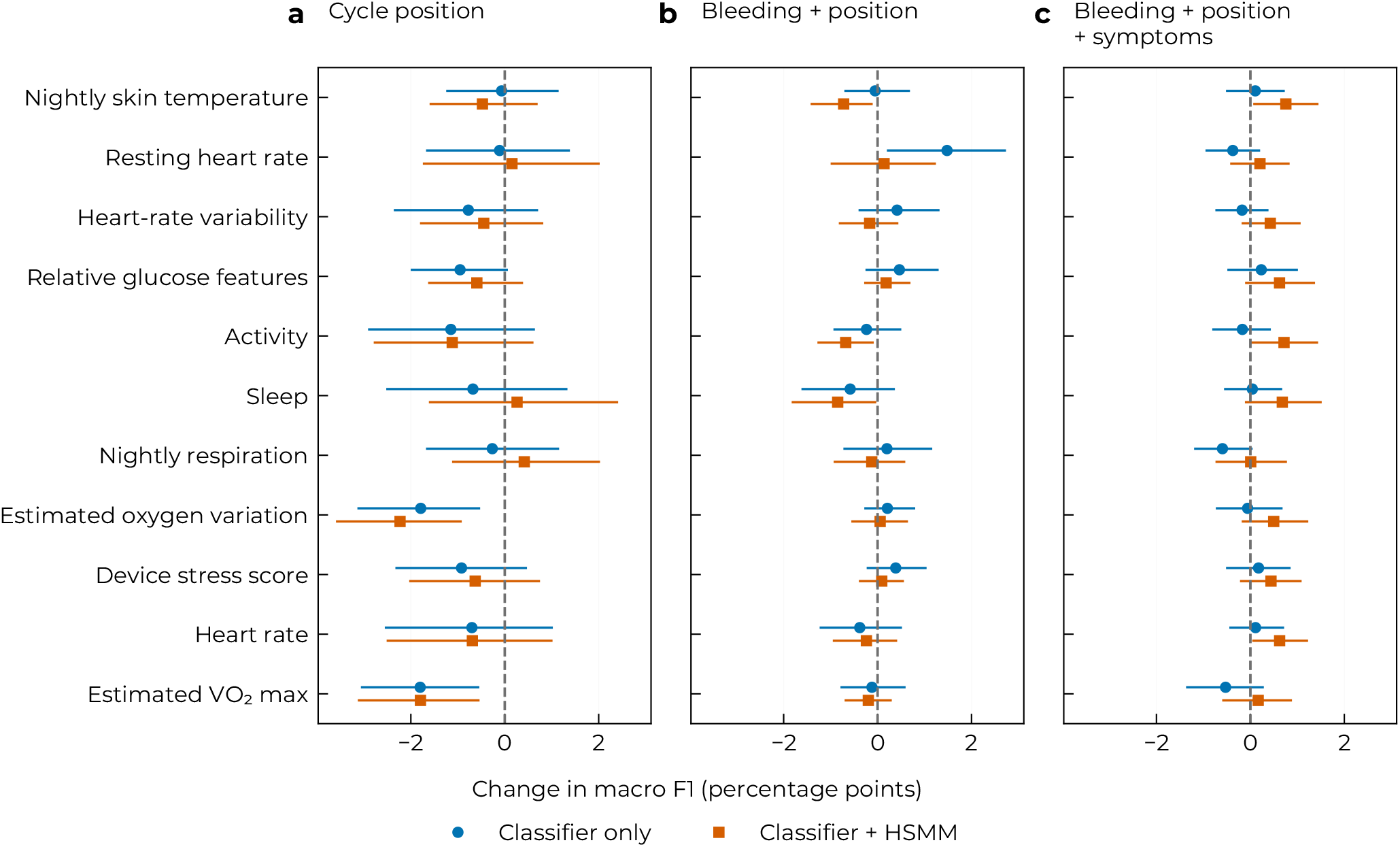
Individual sensor-derived category contributions. Change in macro F1 after retraining CatBoost with each category added to **a**, bleeding-derived position, **b**, bleeding and position, or **c**, bleeding, position and symptoms. Circles denote classifier-only predictions; squares include forward HSMM filtering. Models use history features. Points show mean paired changes across 42 held-out participants, averaging 3 seeds within participant; bars show 95% paired participant-bootstrap intervals (5,000 resamples), without multiplicity adjustment. Positive values favour added inputs; dashed lines indicate no change.

Matched-seed comparisons showed higher macro F1 for both primary learners than for the prior-only baseline, with and without HSMM filtering. Differences from logistic regression, random forest and the multilayer perceptron were uncertain, with all nominal intervals spanning zero (Table S4).

### Effects of temporal representation

Dynamics features improved sensor-only prediction. CatBoost classifier macro F1 increased from 24.11% with history features to 35.17% with dynamics, while forward HSMM performance increased from 30.08% to 43.55% (Figure 3). The largest mean improvements in that comparison occurred in the sensor-only configuration; changes were smaller when bleeding and position were available.

**Figure 3:**
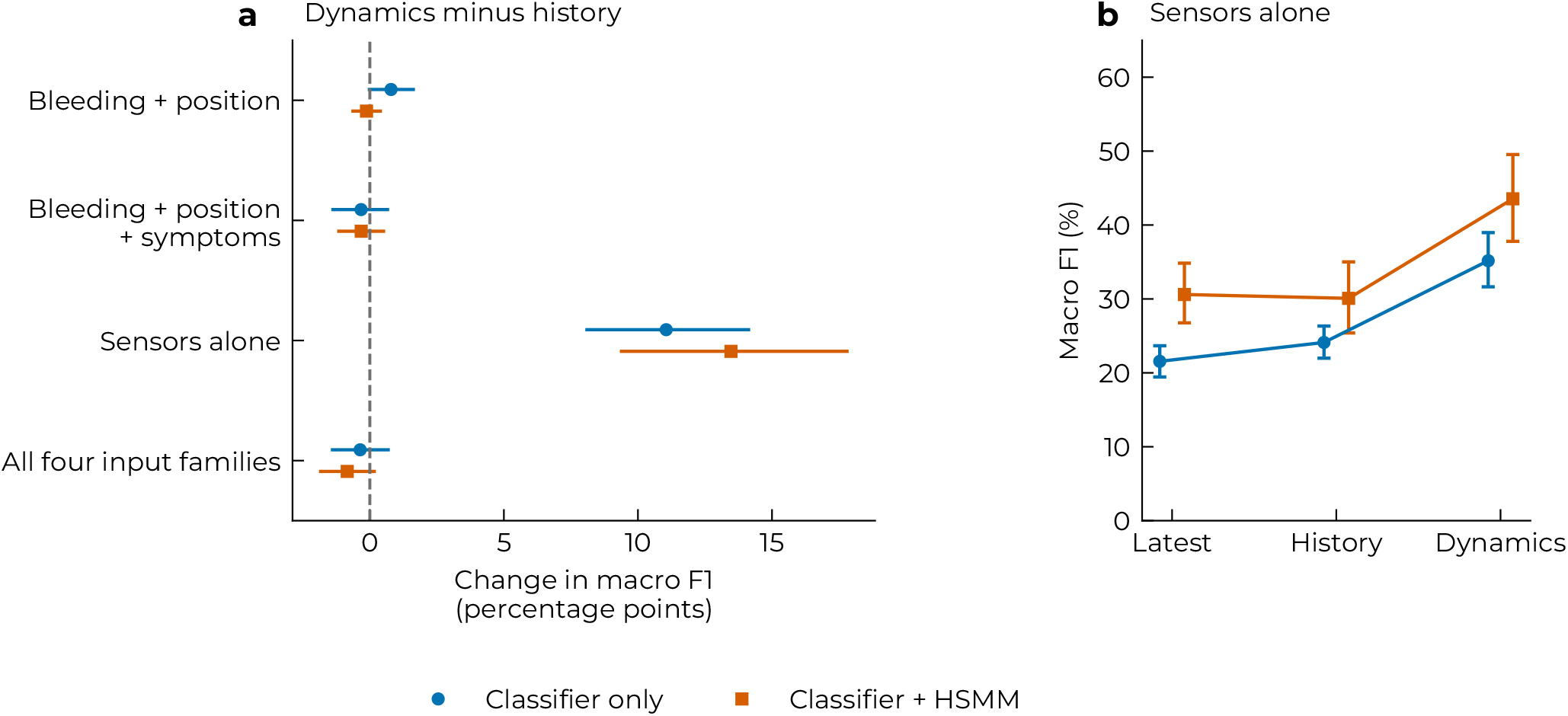
Temporal representation and sensor-only performance. **a**, Change in macro F1 after retraining with dynamics rather than history features for each input combination. Dynamics adds lagged changes, a seven-day trend and deviation from the prior personal mean to the history representation. Positive values favour dynamics. **b**, Sensor-only performance with latest, history and dynamics features. Circles denote classifier-only predictions; squares include forward HSMM filtering. All models use CatBoost. Estimates average 3 seeds within each of 42 held-out participants; bars show nominal 95% participant-bootstrap intervals, paired in a. Sensor inputs use the preceding day; sensor-only models exclude bleeding, position and symptoms.

### Sensitivity to delayed and missing reports

In the scored data, bleeding reports were missing on 15.3% of days and each symptom item on 11.4%; all reports were missing simultaneously on 4.7% of days (Table S9).

Weekly reporting gaps caused larger mean performance losses than one-day delays. For the CatBoost classifier using bleeding and position, delaying reports by one day changed macro F1 by -3.78 percentage points (95% interval [-5.25, -2.39]), compared with -10.28 points ([-13.72, -7.12]) when three consecutive reporting days were removed in each seven-day block. Performance also decreased in configurations including symptoms and sensors; sensor-only predictions were unchanged (Figure 4).

**Figure 4:**
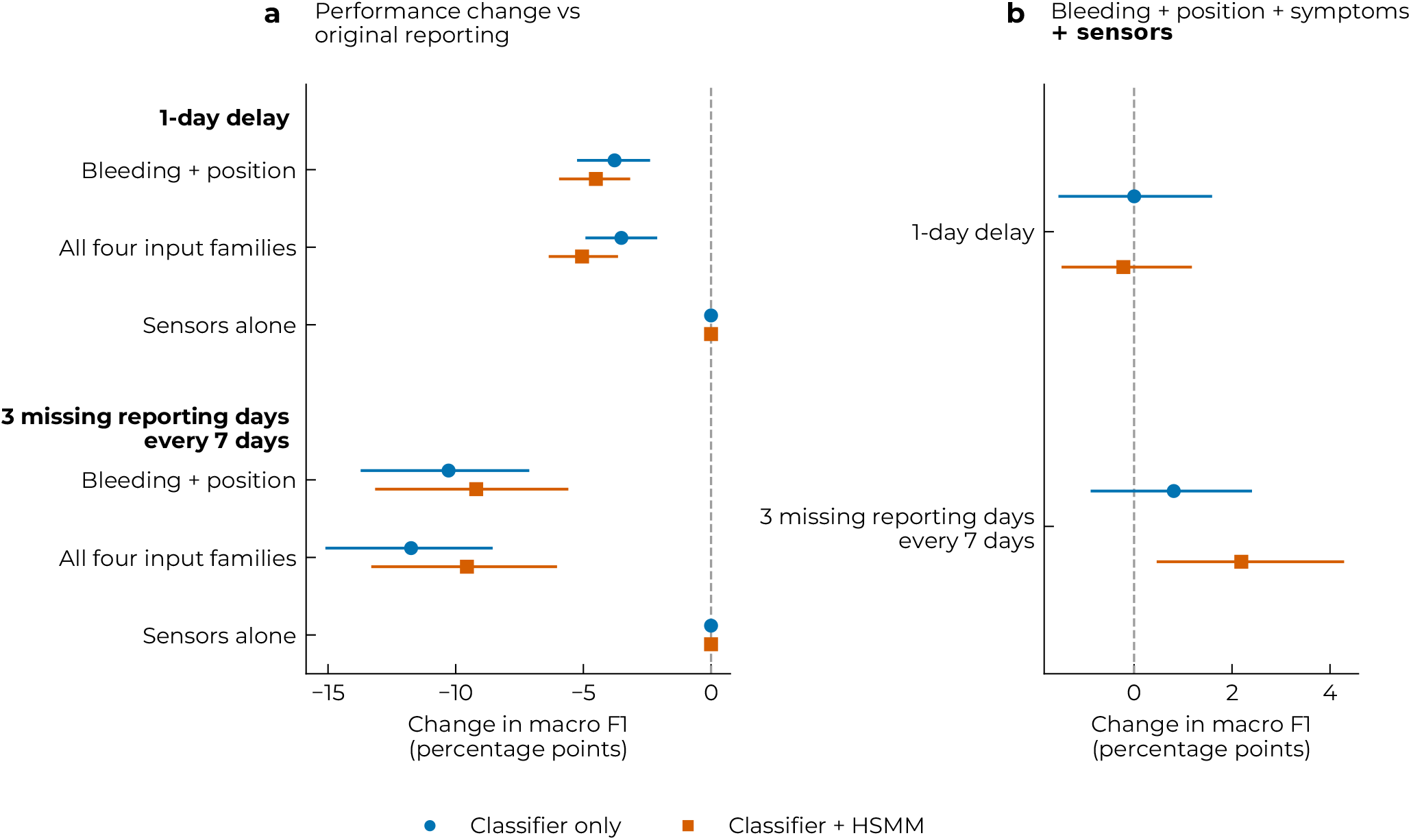
Effects of disrupted reporting and added sensors. **a**, Change in macro F1 with delayed or missing reports versus original reporting, using the same input combination; negative values indicate poorer performance. **b**, Change from adding sensors to bleeding, position and symptoms under the same reporting disruption; positive values favour sensors. Bleeding and symptom reports are delayed by one day or removed on the last three days of every seven-day block from each participant’s start. Gaps are fixed, not random. Models are retrained with scenario-specific histories and reconstructed position; sensor availability is unchanged. Circles denote classifier-only predictions; squares include forward HSMM filtering. All models use CatBoost with history encoding. Points show mean changes across 42 held-out participants; bars show nominal 95% paired participant-bootstrap intervals. Comparisons use matched seed 42; panel b is post-hoc.

In the post-hoc comparison under matched reporting conditions, adding sensors to bleeding, position and symptoms changed classifier macro F1 by +0.00 percentage points (95% interval [-1.54, +1.59]) under one-day delays and +0.81 points ([-0.89, +2.41]) under weekly gaps. Neither comparison established recovery of performance through the added sensor inputs.

### Temporal filtering, phase errors and calibration

Temporal filtering changed transition detection more than average phase-classification performance. For full-input CatBoost with history features, HSMM filtering changed macro F1 from 63.28% to 64.50%. With three-day matching, aggregate transition precision increased from 67.7% to 76.9%, while recall decreased from 73.3% to 72.1% (Table S6). The precision–recall trade-off persisted with one-day matching, while exact-day precision and recall were low across all methods (Figure S9 and Table S6). Participant-level changes varied, with both improvements and decreases following HSMM filtering (Table S5). Timelines illustrate this variation for participants nearest the lower quartile, median and upper quartile of full-input HSMM performance (Figure 5).

**Figure 5:**
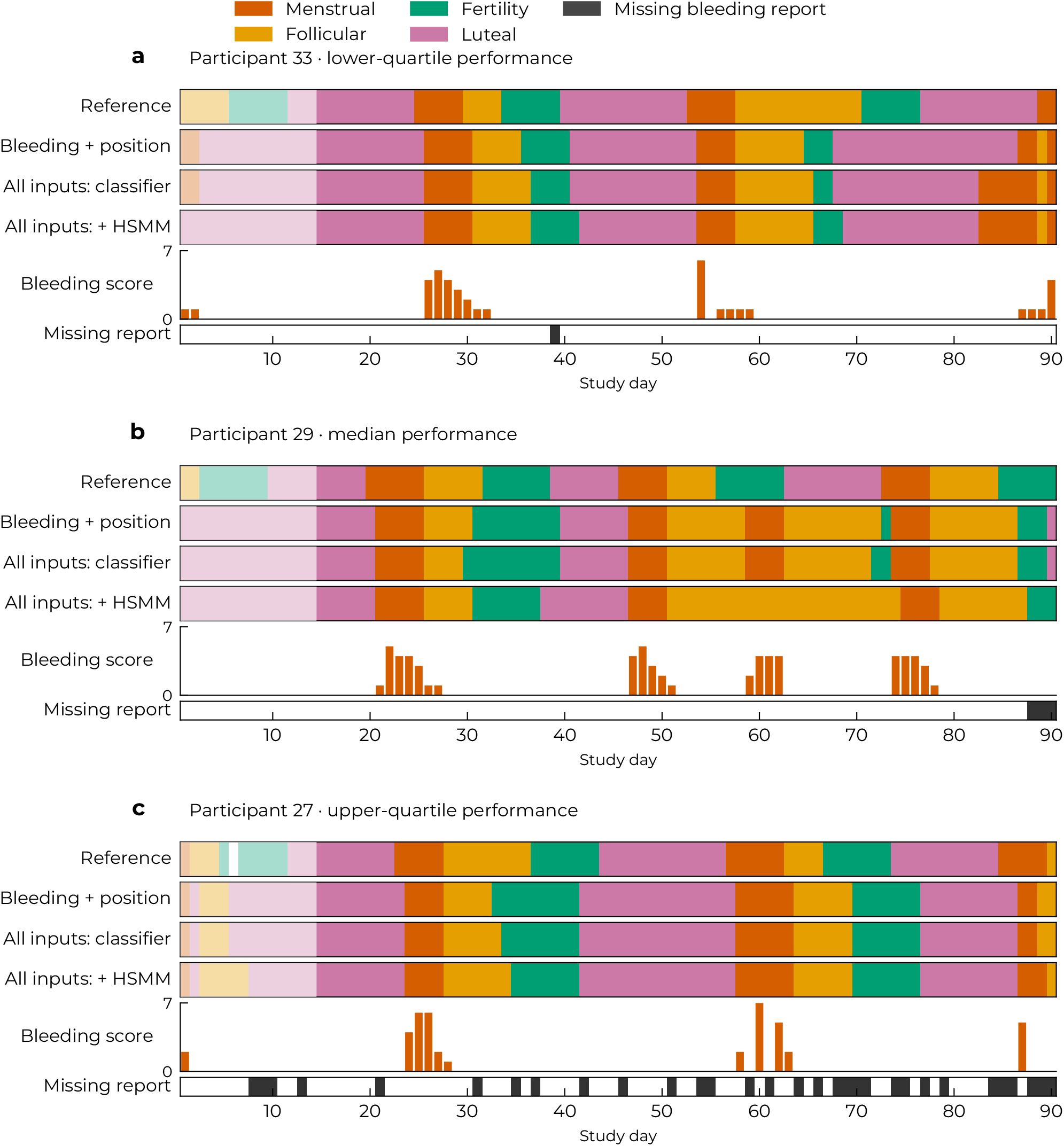
Reference phases, predictions and bleeding. **a–c**, Participants nearest the 25th, 50th and 75th percentiles of macro F1 for full-input CatBoost with HSMM filtering, averaged across 3 seeds. Rows show reference phases and predictions from the bleeding-and-position classifier, full-input classifier and full-input classifier with HSMM filtering. Full inputs comprise bleeding, position, symptoms and sensors. Displayed predictions use held-out models with history encoding and seed 42. Pale regions mark unscored days. Bars show bleeding intensity (0–7); dark cells below identify missing bleeding reports. The horizontal axis shows study day.

Exploratory participant-level analyses showed weak and imprecise associations between reporting completeness and full-input classification performance. Correlations with missing bleeding reports were *−*0.21 for Cat-Boost (95% interval [*−* 0.49, 0.11]) and *−*0.25 for Extra Trees ([*−*0.55, 0.09]). Reporting completeness showed no clear association with gains from adding sensors. For Extra Trees, participants with more missing bleeding reports showed greater gains from HSMM filtering, although this exploratory association was not consistent across both classifiers (Table S7). Only 3 participants had at least three complete reference cycles, providing insufficient support for analysing performance against cycle-length variability. Although 13 participants reported irregular menstruation, the released data did not identify these participants, preventing a comparison by self-reported regularity.

Fertility had the lowest phase-specific F1 and Luteal the highest across the four CatBoost input/output configurations in Table S2. For the full-input classifier, reference Fertility days were classified as Luteal on 36.3% of days and as Follicular on 18.6%, after equal participant averaging (Figure S1). Adding inputs or temporal filtering did not change the ordering of these highest- and lowest-performing phases.

High-confidence HSMM predictions were overconfident. In the highest confidence bin for full-input Cat-Boost, mean confidence was 98.2%, whereas observed accuracy was 72.6% (95% interval [68.4, 76.3]%; Figure S3). Daily probability trajectories show how filtering altered confidence and phase transitions for the same participants used in the timelines (Figure S10).

### Feature attribution and removal

Bleeding had the largest mean absolute grouped SHapley Additive exPlanations (SHAP) attribution, followed by cycle position and symptoms. Among sensor categories, heart rate and activity had the largest grouped attributions (Figure S6). Removing individual sensor categories produced small macro-F1 changes. Several nominal intervals favoured retaining a category, particularly with HSMM filtering, but none remained significant after Holm adjustment (Figure S7). Sensor attribution in the fitted classifier therefore did not establish a robust incremental benefit from retaining a particular category.

## Discussion

Bleeding and bleeding-derived cycle position accounted for much of the phase-classification performance in this cohort. Adding symptoms and wearable measurements produced small, model-dependent changes, with no consistent improvement beyond bleeding and position across the two tree learners. Sensor-only prediction nevertheless improved with features describing changes over time, and models using bleeding and symptoms were sensitive to delayed or missing reports. Together, these findings show that the added value of symptoms and sensors depends on the inputs already available and how changes over time are represented.

Bleeding and cycle position provide a strong starting point for predicting the supplied phase annotations. This result must be interpreted in relation to their construction: menstrual-flow boundaries contribute to the reference phases alongside hormone-based fertile-window estimates [14]. A target established independently of menstrual flow could yield a different comparison.

Previous wearable studies provide evidence of cyclerelated physiological variation. Goodale et al. reported phase-associated changes in wrist skin temperature, heart rate and respiratory rate, and developed a fertile-window classifier [7]. Kilungeja et al. demonstrated menstrual-phase classification from wearable measurements [6]. Such findings are compatible with our sensor-only results. Physiological variation can support prediction without substantially improving a model that already has informative bleeding and position features. Our comparisons extend this work by measuring the additional predictive value of symptoms and sensors against those explicit baselines.

The prediction target also helps explain differences between mcPHASES studies. Xu et al. reported improved next-menses timing using wearable evidence relative to calendar and historical references, with the effects of personal history differing across irregularity profiles [10]. Their post-ovulation timing task asks when the next event will occur, whereas our task assigns a phase to each scored day. Wearable measurements may add more information for predicting the next menstrual onset than for classifying phases partly defined by bleeding, although we did not directly compare these tasks.

Nested feature selection did not reveal a consistent average benefit from adding symptoms and sensors beyond bleeding and position. CatBoost with HSMM filtering showed a nominal sensor benefit when symptoms were already included, but this was not reproduced by Extra Trees and did not remain significant after multiplicity adjustment. The result suggests that input value can depend on the learner and temporal filter, while leaving uncertainty about whether the improvement would generalise. Similarly, prominent SHAP attribution indicates that a fitted classifier uses a feature, while small changes after retraining without it indicate that comparable performance can be obtained from the remaining inputs. Substitutable information is one possible explanation, but these comparisons do not establish redundancy. The confidence intervals leave room for positive effects, but do not identify a reliably beneficial sensor category.

The improvement in sensor-only prediction with dynamics features suggests that changes over time can be more informative than simpler summaries of measurement levels. Kilungeja et al. likewise found that classification performance depended on temporal windowing and phase definitions [6]. Our comparison motivates further study of representations that capture within-person change, particularly when bleeding histories are unavailable. Predictive patterns may reflect measurement availability as well as physiology.

Temporal filtering addressed a separate question: how to combine daily classifier outputs with expected cycle progression. For full-input CatBoost with history encoding, filtering increased transition precision while reducing transition recall, with little change in macro F1 (Figure S9). Thus, fewer false transitions can come at the cost of missed transitions. Whether that trade-off is useful depends on the intended task and the importance of correctly timing phase boundaries.

The reliability curves show overconfidence in parts of the HSMM probability range (Figure S3), indicating that temporal filtering does not ensure well-calibrated probabilities.

For the evaluated target, the results support using bleeding and cycle position as a reference against which additional data collection is assessed. They provide little evidence that collecting every available symptom or sensor stream improves average prediction. This can help focus future studies on the benefit of additional measurements relative to their collection demands, rather than assuming that a larger input set will be more informative.

That comparison must also account for reporting availability. Delays and weekly gaps reduced performance in models using bleeding and symptoms, and wearable inputs did not reliably compensate under the tested schedules. A configuration requiring few input types may still depend on sustained engagement. Prospective studies should therefore evaluate prediction performance together with reporting adherence and participant burden, which were not measured by our simulations or symptom-budget analysis.

Although simulated reporting disruptions reduced performance, observed differences in reporting completeness were not clearly associated with full-input classifier performance across participants. These analyses address different questions: the simulations alter information availability within matched participants, whereas between-participant associations also reflect other individual differences. The wide intervals leave substantial uncertainty about these associations.

The next step is to test whether additional modalities improve tracking against independently established physiological outcomes, using longer records and more diverse cohorts. Longer observation could help distinguish stable individual differences from cycle-related changes and evaluate the role of personal history suggested by Xu et al. [10]. Personalised models could adapt phase-duration priors or classifier parameters to earlier cycles, particularly for people with irregular cycles. Testing this approach requires longer records with participant-linked cycle-regularity information and evaluation on subsequent cycles. The present results do not justify prioritising a particular sensor, but they identify useful evaluation criteria: improvement beyond existing information, robustness to missing reports and well-calibrated prediction probabilities.

Several limitations qualify these findings. The reference phases combine Mira-derived fertile-window estimates with menstrual-flow boundaries. Although the dataset authors report physiological patterns consistent with the assigned phases, independent confirmation of ovulation was not reported for this cohort [14]. Agreement with these annotations therefore does not establish accuracy against an independent physiological reference, and their dependence on bleeding may favour bleeding-based predictors.

The cohort comprises 42 young adults from the Greater Toronto Area, most identifying as East or South-east Asian, and excludes hormonal contraceptive users [14]. Evaluation on held-out participants assesses generalisation within this cohort. Applicability to other populations and variation in performance by cycle regularity or reproductive conditions remain uncertain.

The analyses are exploratory, and modelling choices were informed by prior investigation of this cohort. Nested validation separates participants during fitting and tuning, but does not remove the possibility that prior exploration influenced the research design. The small sample limits precision, particularly for phasespecific comparisons. Uncertain differences between input combinations should not be interpreted as evidence of equivalent performance.

Predictors used only information available by the prediction cutoff, but daily aggregation, questionnaire timing and the initial warmup period may differ from prospective use. Simulated reporting gaps may not reflect natural missingness, and evaluation on unseen participants does not assess personalised models trained on an individual’s earlier cycles. Device-derived scores may reflect proprietary algorithms as well as physiology.

Finally, the evaluated models, feature-selection strategy and temporal representations cover a limited range of approaches. Other modelling choices may change the estimated benefit of symptoms and sensors. Longer records and independent cohorts are needed to assess whether the findings extend beyond the present study.

In this mcPHASES analysis, symptoms and wearable measurements showed little consistent improvement in daily menstrual phase classification beyond bleeding and bleeding-derived cycle position. Sensor-only prediction improved when features captured changes and trends over time, whereas wearable inputs did not reliably compensate for the simulated reporting disruptions. These findings support evaluating additional measurements against explicit bleeding and cycle-position baselines to establish their incremental predictive value. Because menstrual flow also contributes to the reference phase definitions, the findings describe prediction of these annotations rather than independently established physiological phases. Fertility remained the least accurately classified phase by F1, and these results do not establish reliability for fertility or contraceptive decisions.

## Methods

### Dataset and reference phases

We performed a secondary analysis of de-identified data from mcPHASES version 1.0.0 [13, 14]. The source study recruited 50 menstruating adults in the Greater Toronto Area, of whom 42 consented to release their data for future research. Diabetes and hormonal therapy or contraception within three months before enrolment were exclusion criteria. The released cohort had a mean age of 20.9 years (standard deviation 2.8); 13 participants reported irregular menstruation (Table 2). The original study was approved by the University of Toronto Research Ethics Board (Protocol 41568), with written informed consent for participation and separate consent for data release and future use. Secondary use is governed by the mcPHASES data use agreement [14].

**Table 2:** Characteristics of the 42 participants in the released 2022 cohort, as reported by the dataset authors [14]. Values are counts unless otherwise stated.

| Characteristic | Value |
|---|---|
| Age, years (mean ± SD) | 20.9 ± 2.8 |
| Self-reported menstruation |  |
| Regular | 29 |
| Irregular | 13 |
| Ethnicity |  |
| East Asian | 14 |
| Southeast Asian | 11 |
| Caucasian | 9 |
| Middle Eastern | 5 |
| African | 1 |
| Indo-Caribbean | 1 |
| Latina | 1 |

We used the 2022 collection interval because it included all input families needed for the modality comparisons. The 2024 follow-up omitted glucose monitoring and made symptom diaries optional. All 42 participants were retained, including those with prolonged reporting gaps; no cycles were excluded on the basis of reporting completeness. The daily grid contains 3,698 participantdays. We used the first 14 calendar days of each participant’s record as a warmup for constructing historical features, including 14-day summaries, and excluded those days from scoring. The same warmup was applied to all input configurations to keep the evaluation period consistent. Days with missing reference labels were not scored. This leaves 3,110 scored days (Figure 6); missing measurements within the historical windows were handled as described in section “Input families and feature construction”.

**Figure 6:**
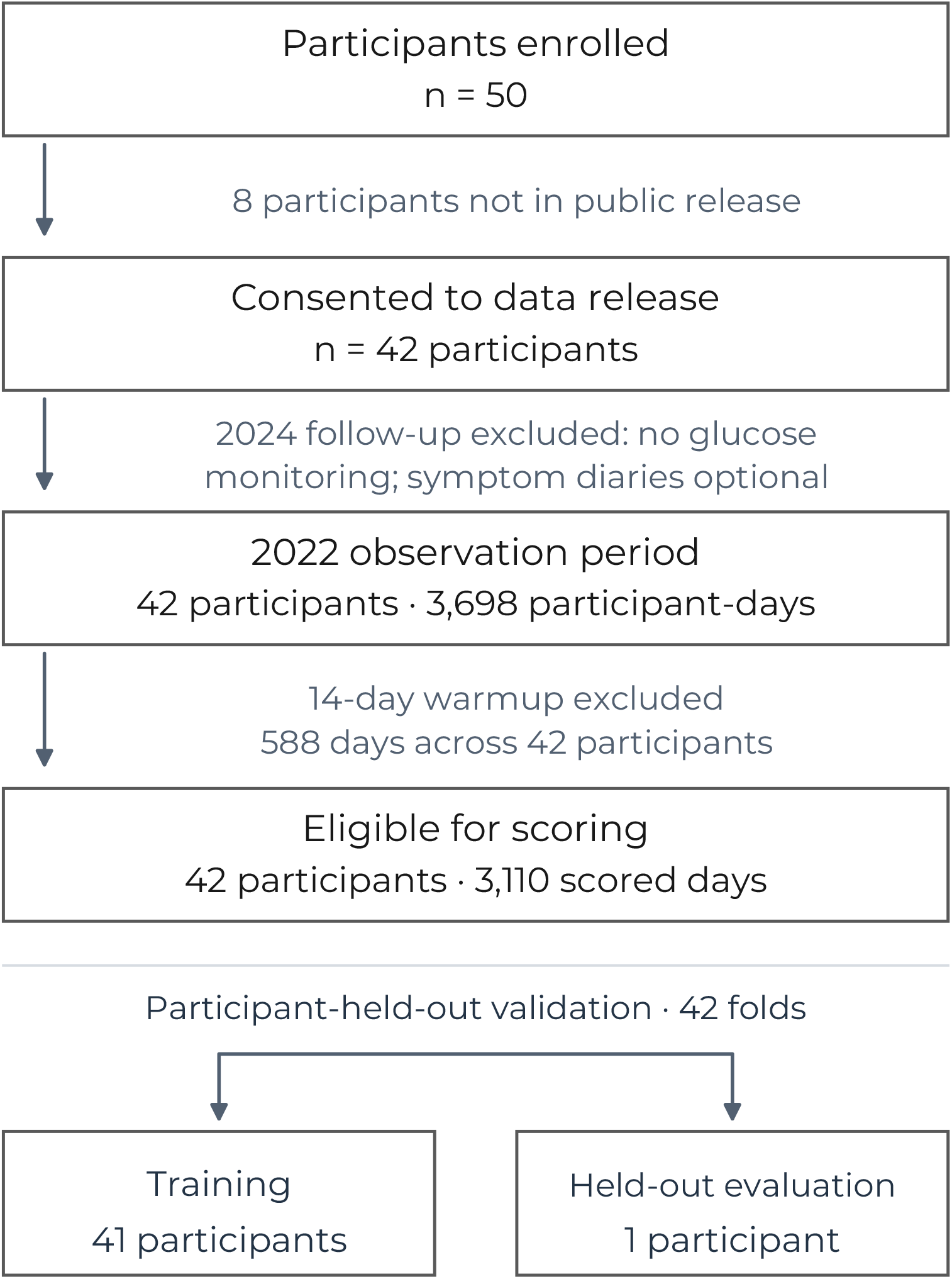
Participant flow and evaluation cohort. Of 50 enrolled participants, 42 consented to data release. The 2022 analysis includes 3,698 participant-days; excluding the 14-day warmup leaves 3,110 scored days. Each of 42 outer folds uses 41 participants for training and one for held-out evaluation.

The four targets are the dataset’s Menstrual, Follicular, Fertility and Luteal reference labels. Menstrual days are defined by menstrual flow, Follicular days span the interval between flow and the fertile window, Fertility corresponds to the estimated fertile window, and Luteal days span the interval between that window and the next menstrual flow. These definitions combine Mira’s hormone-based fertile-window estimates with menstrual-flow boundaries [14]. Fertility denotes an estimated fertile window rather than a confirmed ovulation day. Hormone measurements and phase labels were excluded from predictors. We included self-reported bleeding as a readily obtainable input that does not require hormone testing, and as a practical baseline for evaluating the added value of symptoms and sensors. However, menstrual-flow reports also contributed to the reference phase definitions, which should be considered when interpreting bleeding-based performance.

Observation coverage and reference durations are summarised in Table S8. 40 participants contributed 90 observation days, with the remaining 2 contributing 38 and 60 days. We identified 76 complete reference cycles across 41 participants, with a median length of 29 days (interquartile range 27–33). Complete cycles required two observed menstrual onsets and an uninterrupted sequence of all four reference phases. An observed onset required a non-Menstrual label on the preceding day; complete phase segments required observed neighbouring labels at both boundaries. Reporting gaps were consecutive missing-report runs on the full grid, including observation-boundary runs whose full lengths may be censored. Zero or Not at all responses counted as observed, and reporting gaps alone did not exclude a reference cycle. Observation-boundary fragments and phase segments truncated by boundaries or missing labels were counted separately. Descriptive durations use the full observation period, including warmup.

### Prediction task and input availability

We estimate the current day’s menstrual phase after the daily questionnaire is available. Report inputs are available at this prediction cutoff, while sensor inputs use the preceding day’s summaries. This one-day delay ensures that sensor summaries cover a completed day. Historical features and temporal filtering use observations available through the prediction cutoff. Missing reports are retained as missing values.

### Input families and feature construction

We group non-hormonal inputs into four families: bleeding, bleeding-derived cycle position, non-flow symptoms and sensors (Table 3). Symptoms and sensors are defined separately from bleeding and position so that each family’s contribution can be evaluated. Position features are derived from bleeding reports. Hormonal assays, reference phases and demographic identifiers are excluded from predictive inputs.

**Table 3:** Input families used for phase estimation. Counts exclude additional availability indicators and temporal-history features. Feature definitions are provided in section “Input families and feature construction“.

| Family | Information represented | Base features |
|---|---|---|
| Position | Time since detected bleeding onset, represented as elapsed time and position within a fixed 28-day reference cycle | 4 |
| Bleeding | Self-reported menstrual-flow volume and colour | 10 |
| Symptoms | Self-reported appetite, exercise level, headaches, cramps, sore breasts, fatigue, sleep issues, mood swings, stress, food cravings, indigestion and bloating | 12 |
| Sensors | Daily measurements, device-derived scores and recording-coverage summaries for nightly skin temperature, resting heart rate, heart-rate variability (RMSSD), relative glucose features, activity, sleep, nightly respiration, estimated oxygen variation, device stress score, heart rate and estimated VO<sub>2</sub> max | 43 |

*Note:* Position is derived from bleeding reports; the fixed reference cycle does not estimate an individual’s cycle length.

Sensor inputs include physiological summaries, device-generated scores and recording coverage. Temperature represents nightly skin temperature; heart-rate variability uses log-transformed daily median root mean square of successive differences between interbeat intervals (RMSSD). Glucose inputs comprised relative variability and change measures rather than absolute concentrations. Estimated oxygen variation is analysed on its exported signal scale, not as oxygen saturation. Fitness is represented by the device-provided VO_2_-max estimate, aggregated by the median within each participant-day. Sleep combines duration and efficiency measures with a device sleep score; device stress is a separate composite score from self-reported stress. Table S10 provides a compact dictionary of the source tables, measurements and units.

Processing was performed separately for each participant and study interval. Exact duplicate records were removed. Repeated timestamps were combined by the median of valid values for glucose, heart rate, oxygen variation and activity streams. Sleep, temperature and five-minute HRV intervals were assigned to their end day before applying the sensor delay. Invalid records were excluded; daily value summaries remained missing when coverage was insufficient, while recording-coverage features were retained.

Nightly skin-temperature records required positive sample counts, finite values and sleep duration in (0, 24] hours; daily temperatures were weighted by sample counts. Resting heart rate and estimated VO_2_ max used daily medians of finite positive values. HRV required finite positive RMSSD and coverage of at least 0.8; its daily median required at least 12 intervals and was transformed using ln(1 + RMSSD). Glucose summaries required finite positive values, at least 144 distinct timestamps and at least 12 hours between the first and last timestamp, including the final five-minute sampling interval. The five glucose features were SD/mean, interquartile range/median, 90th/10th percentile ratio, daily mean divided by the preceding seven-day mean (at least three observed days), and observed fraction (sample count/288, capped at one).

Activity used four intensity-minute and four heartrate-zone values, each set finite and nonnegative with a total of 1–1440 minutes, aggregated by daily median. Steps, distance, calories and altitude gain used sums over valid nonnegative records plus observed-minute counts.

Sleep used the longest valid main episode ending that day, requiring elapsed time in (0, 24] hours, recorded duration of 1–86,400,000 ms, asleep/awake minutes of 0– 1440, time in bed of 1–1440 minutes and efficiency of 0–100; the daily sleep score was the median of values from 1 to 100. Nightly respiration used medians of positive full-night rates and nonnegative SDs, requiring positive signal-to-noise. Oxygen variation retained signed values; its minute medians required 120 observed minutes for daily mean, SD, median and interquartile range. Heart-rate summaries used the same four statistics on positive minute-level values, requiring 360 observed minutes. Both streams also contributed observed-minute counts. Device stress used median scores from 1 to 100 with READY status and a false calculation-failed flag.

RMSSD and estimated VO_2_ max units follow the Fitbit/Google Health API documentation. Although the dataset README specifies mmol/L for glucose, exported values span 2.2–15.5 in 40 participants and 40– 253 in 2, without per-record units. Dimensionless glucose ratios avoid assigning absolute concentrations, but ratios to prior days assume consistent scaling within participant.

Flow volume is encoded on an ordinal scale from 0 (Not at all), 1 (Spotting/Very Light), 2 (Light), 3 (Some-what Light), 4 (Moderate), 5 (Somewhat Heavy), 6 (Heavy), to 7 (Very Heavy). Flow colour is represented by nine one-hot categories, including separate categories for Other and Not at all.

A bleeding onset occurs when volume is at least 2 after two low-flow days. Let *d* be days since the detected onset (days since bleed (DSB)). Position consists of *d*, sin(2*πd/*28), cos(2*πd/*28) and min(*d/*28, 2). The sine and cosine terms provide a repeating representation using a fixed 28-day reference period. The clipped term provides a bounded measure of elapsed time, reaching its maximum at 56 days, while *d* retains the unbounded elapsed-day value. The reference period and cap are fixed modelling choices rather than participant-specific estimates of cycle duration. Report-delay and gap scenarios reconstruct position from the available reports.

Wearable measurements vary across the menstrual cycle [7], and previous wearable-based phase classifiers have incorporated slopes and changes within temporal windows [6]. We compare three temporal representations to examine whether predictive performance depends on explicitly representing changes over time. The latest encoding contains available base values and their availability flags. History adds 7-and 14-day means, sample standard deviations and observed fractions for non-position inputs. These fixed windows summarize measurements at weekly and fortnightly timescales, providing shorter and longer temporal context. Window lengths were not tuned and do not represent individual phase boundaries. Dynamics further adds differences from 1, 3 and 7 days earlier, a 7-day regression slope, and deviation from the participant’s expanding prior mean. Rolling summaries include the current available value, which for sensors represents the preceding day. Means, standard deviations and slopes require at least three observed values; observed fractions use the available window from its first row. The expanding baseline uses only prior values and requires seven observations. Missing derived values are retained until fold-specific preprocessing.

These transformations produce 138, 528 and 853 candidate columns for the full-input latest, history and dynamics representations, respectively. History offers 88 columns for bleeding and position and 184 when symptoms are added. Within each training fold, preprocessing removes constant columns and redundant columns with absolute Pearson correlation greater than 0.95. CatBoost handles missing numerical values natively [15]; Extra Trees uses median imputation fitted on the training participants. These steps are repeated within inner training folds and then on outer training participants. Retained columns therefore vary by fold; the generated feature inventory and saved fold manifests provide exact names and selections.

### Models and temporal filtering

CatBoost [16] and Extra Trees [17] provide two nonlinear tabular learners with different tree-building procedures. Comparing them tests whether incremental modality findings depend on the learner. CatBoost candidates use 200 trees, depth 3 or 5, learning rate 0.05 and L2 regularisation 3. Extra Trees candidates use 250 trees, a minimum leaf size of 3 or 10, and consider all available features at each split.

Daily classifiers do not explicitly account for the ordered progression and duration of menstrual phases. We therefore evaluated whether incorporating these temporal patterns improves phase estimation beyond daily classifier predictions. We compared the classifier alone with a hidden Markov model (HMM), which represents phase order and persistence through transition probabilities, and a HSMM, which additionally models phase-duration distributions explicitly [18]. This comparison tests whether temporal structure improves prediction and whether explicit duration modelling provides further benefit. Both models used forward filtering, so each day’s prediction depended only on observations available up to that day. Figure 7 summarises the prediction pipeline. Both use the cyclic order Menstrual*→* Follicular *→*Fertility*→* Luteal. For each phase, duration probabilities were estimated from complete training segments lasting 1–60 days. We applied weak smoothing to avoid assigning zero probability to durations not observed in training. Complete segments required observed labels at both bounding transitions and eligible days throughout; segments truncated by recording boundaries or missing labels were excluded. For phase *s* and duration *d*, probabilities were (*n*_*s,d*_ + *q*_*d*_)*/*(*N*_*s*_ + 1), where *n*_*s,d*_ is the training count and *N*_*s*_ the total eligible segment count. The smoothing distribution *q*_*d*_ was geometric with success probability 0.1, with mass for durations of 60 days or longer pooled at day 60. Its total weight equalled one segment per phase. The common smoothing distribution did not impose a common fitted mean duration. HMM transition probabilities counted same-phase and next-phase transitions between consecutive eligible labelled days, adding one count to each allowed transition before normalisation.

**Figure 7:**
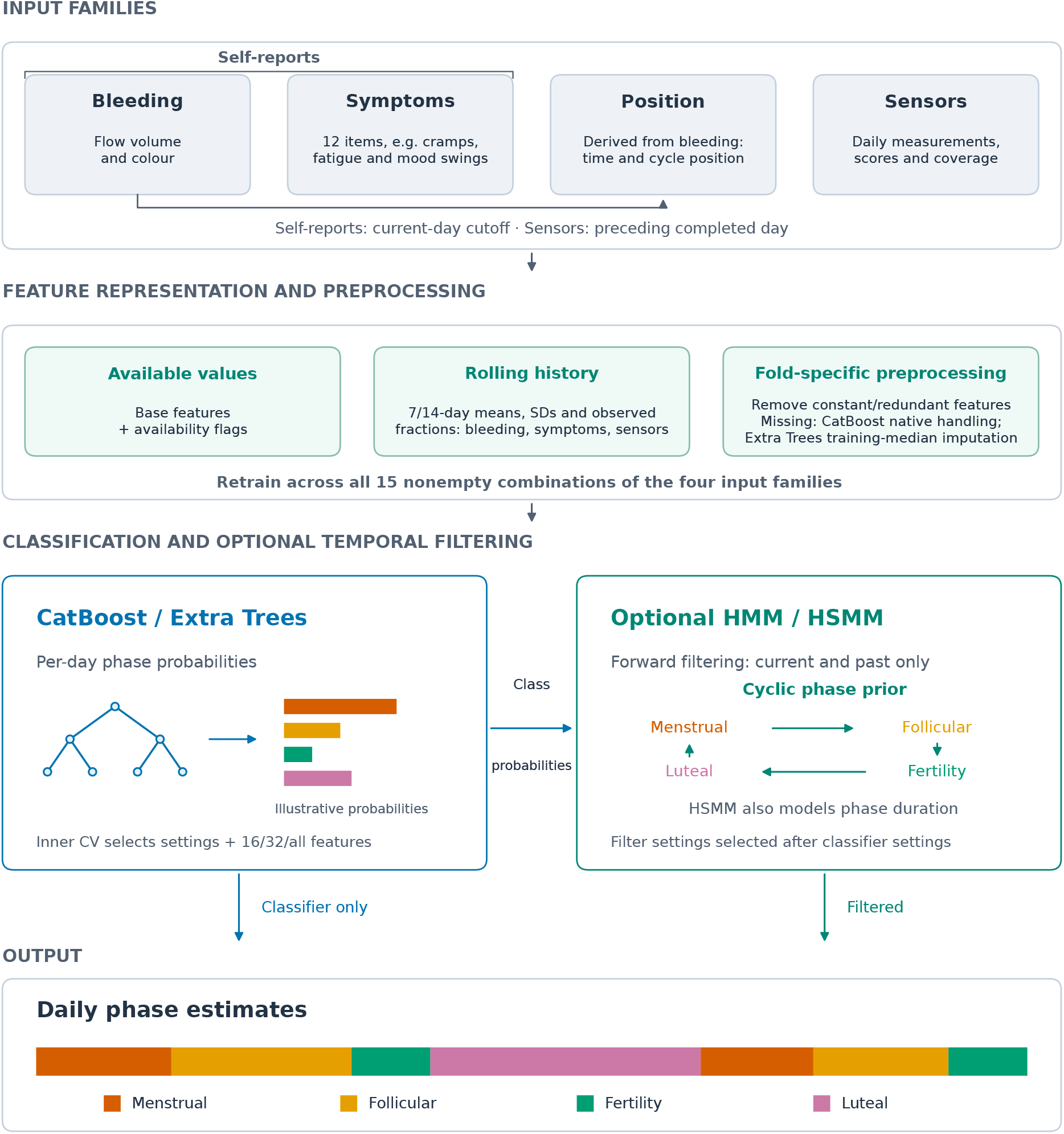
Prediction pipeline for menstrual phase estimation. Self-reports comprise bleeding and symptoms; position is derived from bleeding reports. Each input combination is evaluated with CatBoost and Extra Trees using classifier-only predictions or forward HMM/HSMM filtering. The primary history representation adds 7- and 14-day summaries to bleeding, symptoms and sensor inputs. Within each outer training set, participant-level cross-validation jointly selects classifier settings and 16, 32 or all features, then selects filter settings with the classifier fixed. Preprocessing and temporal priors use training participants only. Selected models are refitted on the outer training participants and evaluated on the held-out participant. The temporal filters use a cyclic phase prior, with explicit durations in the HSMM; pointwise phase predictions can still skip phases. Probabilities and phase sequences are schematic.

For classifier probability *p*_*t*_(*s*), the emission weight is max(*p*_*t*_(*s*), 10^*−*12^)^*α*^, with *α ∈ {*0.25, 1, 2 }. A reset probability of 0 or 0.05 permits recovery from an incorrect latent state. Forward filtering updates the state distribution using observations available through day *t*. The selected reset probability remains fixed during held-out evaluation; bleeding events and reference labels do not trigger resets. Pointwise maximum-probability outputs can skip phases despite the cyclic latent-state prior; we quantify these noncyclic transitions in the evaluation.

### Participant-level validation and model selection

Given the small cohort, we use 42 outer leave-one-subject-out (LOSO) folds to retain 41 participants for training while evaluating each participant in turn. All days from a participant remain together because repeated observations share individual characteristics and temporal dependencies. This separation evaluates generalisation to unseen participants.

Within each outer training set, 3-fold participant-level cross-validation jointly selects classifier settings and a feature budget of 16, 32 or all available columns. Reduced subsets are ranked by an Extra Trees model fitted only to the corresponding training participants; the full-feature option retains the learner’s usual preprocessing. We calculate macro F1 separately for each validation participant and select the combination with the highest mean score. This procedure is applied throughout the experiment matrix, except for the prior-only base-line, which has no input features. Ranking uses training-median imputation and an Extra Trees classifier with 200 trees, a minimum of five samples per leaf, square-root feature sampling and a fixed seed. Ties favour the full feature set, followed by the earlier classifier candidate. In the symptom-budget analysis, questionnaire items are selected before engineered columns.

We then keep the selected classifier configuration fixed and select the HMM and HSMM settings separately using the same inner validation predictions. For each inner fold, transition and duration patterns are estimated only from its training participants. Candidate filter settings are compared by mean participant-level macro F1.

Finally, we recompute any selected feature ranking, fit the classifier and estimate the temporal priors using all 41 outer training participants, then evaluate the classifier alone and with temporal filtering on the held-out participant. Preprocessing is fitted separately within each training fold. We repeat each primary configuration with seeds 17, 42 and 73.

#### Input comparisons and sensitivity analyses

We evaluate bleeding, position, symptoms and sensors individually and in every combination (15 input configurations), using CatBoost and Extra Trees on the same scored participant-days. Comparisons between models retrained with and without each input family quantify its added predictive value. The main comparisons add symptoms to bleeding and position, then sensors to bleeding, position and symptoms. Individual sensor categories are added to three baselines: position alone; bleeding and position; and bleeding, position and symptoms. We also remove individual sensor categories from the full-input model and retrain to assess their contribution when the remaining inputs are available. We selected comparison models with different levels of complexity: a prior-only baseline without input features, logistic regression as a linear comparator, and random forest and a multilayer perceptron as alternative nonlinear approaches. These comparisons assess whether predictive performance depends on the choice of learning algorithm (Table S3).

The main comparisons use history features, which add 7- and 14-day summaries to the available measurements. Additional analyses use latest values or add changes and trends to test whether the findings depend on feature representation. Models using only measurement-availability indicators and observed fractions test whether recording patterns themselves predict phase.

We simulate two reporting disruptions: a one-day delay in the availability of bleeding and symptom reports, and missing reports on the last three days of each sevenday block in a participant’s record. Historical features and bleeding-derived position are recalculated from the reports available under each scenario, while sensor inputs remain unchanged. Models are trained and evaluated under the same reporting scenario; each comparison uses a single, matched random seed for the modified and observed reporting conditions. An additional post-hoc comparison tests whether adding sensors to bleeding, position and symptoms improves prediction under the same reporting disruption.

We examine the effect of reducing the number of symptom questions by comparing models using 0, 2, 4, 6, 8 or all 12 symptom items alongside bleeding and position. Symptoms are ranked by their association with the reference phases using training data only, separately within each training fold. All features derived from unselected symptom items are excluded.

To examine sensitivity to feature selection, we compare the nested-selection pipeline with the otherwise matched pipeline using all available features. The comparison evaluates the selected ranking strategy using the same participant splits, scored days and random seeds (Figure S8). This exploratory comparison was developed after earlier analyses of the cohort and a pilot. Classifier-only modality increments are compared using paired participant-bootstrap intervals without additional hypothesis tests.

### Performance measures and statistical analysis

The primary outcome was macro F1, averaged across 3 random seeds within each participant and then equally across participants. Secondary outcomes included accuracy, phase-specific precision, recall and F1, calibration, and phase-transition performance.

Nominal 95% intervals use 5,000 participant bootstrap resamples, paired for differences. Two-sided paired signflip tests use 49,999 randomisations. Holm adjustment is applied separately across the specified macro-F1 contrasts and the secondary-endpoint contrasts. Confidence intervals are not multiplicity-adjusted.

In a post-hoc analysis, we compared each full-input primary learner with the prior-only, logistic-regression, random-forest and multilayer-perceptron baselines, separately for classifier and HSMM outputs. These comparisons use seed 42 on both sides, matching the comparator runs, with the same participants and scored days. Paired macro-F1 differences use participant-bootstrap intervals and sign-flip tests as above, with Holm adjustment across the 16 baseline-model comparisons (Table S4).

Transitions are matched one-to-one by destination phase. We evaluate exact-day matching and tolerances of one and three days, using one-day matching in Figure S9; the three-day tolerance assesses sensitivity to a wider matching window. The same saved predictions are used at every tolerance. Counts are averaged across seeds within participant, then summed across participants before calculating precision and recall. Table S6 reports all tolerances and timing errors. We report missed and spurious transitions as well as timing error among matched transitions. Transitions across missing-label gaps are excluded. Aggregate transition counts are descriptive. Reliability curves assess the classifier and temporal-filter probabilities directly, without temperature scaling (Figure S3).

We explored participant-level associations between reporting completeness and held-out performance using Spearman correlations with 5,000 paired participant-bootstrap resamples and nominal 95% percentile intervals. Outcomes were full-input classifier macro F1, the change after adding sensors to bleeding, position and symptoms, and the change after HSMM filtering of the full-input classifier. Each participant’s scores were averaged across seeds before calculating differences. Missingness was measured on scored days; symptom missingness was averaged across the 12 items. These posthoc analyses used history encoding and observed reports; secondary descriptors were the longest missing-bleeding run, scored-day count, complete reference-cycle count and cycle-length SD. Unscored or nonconsecutive days broke a missing-report run. Cycle-length variability required at least three complete cycles. Before inspecting associations, we required at least ten supported participants and at least five participants differing from the modal descriptor value; cycle-length variability and scored-day count did not meet these criteria. Spearman correlations were recomputed within each resample, without hypothesis tests or multiplicity adjustment. Inference for these associations and the baseline-model contrasts is conditional on fitted predictions and excludes uncertainty from model refitting and overlapping training sets; the matched single-seed baseline contrasts also exclude variation across training seeds.

### Model interpretation and reproducibility

Tree SHAP [19] quantifies feature contributions to the raw class scores of full-input CatBoost models with history features; it does not explain the subsequent temporal filtering. The analysis covers 126 fitted models, one for each of 42 held-out participants and 3 random seeds. Reconstructed models are verified to reproduce the saved held-out probabilities exactly before calculating SHAP values. For each model, absolute SHAP values are averaged across the held-out participant’s scored days and the four class scores, then across seeds within participant and equally across participants. Features excluded during training receive zero attribution for that fold. Category attribution sums signed feature contributions within category before taking magnitude and applying the same averaging procedure. These attributions describe how the fitted classifier uses its inputs; the retrained comparisons in section “Input comparisons and sensitivity analyses” assess their incremental predictive value.

The analysis package provides a command to run the analyses and export the figures and tables using the supplied dependency versions. The accompanying notebook reads and displays the generated outputs. Participant data are obtained separately under the mcPHASES access conditions (see Data Availability and Code Availability).

## Data Availability

The data analysed in this study are available from the mcPHASES dataset (version 1.0.0) on PhysioNet at ht tps://doi.org/10.13026/zx6a-2c81 [13, 20]. Access is restricted to registered users who sign the dataset’s data use agreement. The participant-level data are not redistributed with the analysis code.

## Code Availability

The analysis code and reproduction notebook are publicly available at https://github.com/MyelinZ/mcphases-classification; the version accompanying this manuscript is revision `2d0876e`.

## Acknowledgements

Not applicable.

## Author Contributions

B.S., S.G. and Z.T. conceived and designed the study. B.S. and M.E. developed the methodology, curated the data and implemented the software. B.S. and Z.T. performed the formal analysis. B.S. prepared the figures. B.S. and Z.T. wrote the original draft of the manuscript. Z.T. and S.G. administered the project. D.K., R.S. and Z.T. supervised the research. All surviving authors reviewed and approved the final manuscript.

