## Supplementary Information for "Evaluating symptoms and sensors for menstrual phase estimation beyond bleeding and cycle position"

#### Contents


**Additional classification performance** **2**

**Input contributions and sensitivity analyses** **8**

**Temporal filtering and participant variation** **13**

**Cohort summaries and feature dictionary** **17**

**Tables**

S1: Classification accuracy 2
S2: Phase-specific performance 3
S3: Additional classifiers 3
S4: Paired baseline comparisons 4
S5: Participant variation 13
S6: Transition-timing sensitivity 14
S7: Participant associations 16
S8: Observation coverage and durations 17
S9: Reporting completeness 18
S10: Feature dictionary 19

**Figures**

S1: Phase-classification confusion 5
S2: All input combinations 6
S3: Calibration 7
S4: Phase-specific input contributions 8
S5: Phase-specific sensor contributions 9
S6: Grouped feature attribution 10
S7: Sensor-category removals 11
S8: Feature-selection sensitivity 12
S9: Temporal filtering 13
S10: Daily phase probabilities 15

### Additional classification performance

Tables S1–S4 report accuracy, phase-specific performance and comparisons between learning algorithms. Figures S1–S3 show phase errors, performance across all input combinations and probability calibration.

Table S1: Classification accuracy by input combination. Values are participant-average accuracy (%) with nominal 95% participant-bootstrap confidence intervals in parentheses. 3 seeds are averaged within each of 42 participants; all models use history encoding. Bold indicates the highest displayed mean within each model and prediction method, including ties; it does not imply a statistically significant difference. All four input families comprise bleeding, position, symptoms and sensors.

| Inputs | Classifier only | Classifier + HSMM |
|---|---|---|
| **CatBoost** | | |
| Cycle position | 59.7 (54.7, 64.5) | 59.1 (54.3, 63.9) |
| Bleeding + position | 66.7 (62.7, 70.5) | 67.9 (64.2, 71.4) |
| Bleeding + position + symptoms | 66.8 (62.8, 70.7) | 67.4 (63.5, 71.1) |
| All four input families | **67.0** (63.4, 70.7) | **68.3** (64.7, 71.9) |
| **Extra Trees** | | |
| Cycle position | 58.7 (53.8, 63.5) | 59.5 (54.4, 64.4) |
| Bleeding + position | **67.9** (64.3, 71.3) | 67.5 (64.0, 71.0) |
| Bleeding + position + symptoms | 67.5 (63.8, 71.1) | **67.6** (63.9, 71.1) |
| All four input families | 67.8 (64.4, 71.2) | **67.6** (64.1, 71.1) |

Table S2: Phase-specific performance of CatBoost. Values are participant-average precision, recall and F1 (%) with nominal 95% participant-bootstrap confidence intervals in parentheses. CatBoost uses history encoding, with 3 seeds averaged within participant. Undefined participant-level metrics are excluded from the corresponding averages.

**CatBoost with history features**

| Phase | Precision | Recall | F1 |
|---|---|---|---|
| **Bleeding + position: classifier only** | | | |
| Menstrual | 81.3 (75.7, 86.7) | 63.9 (57.7, 69.5) | 68.8 (62.7, 74.1) |
| Follicular | 69.3 (63.3, 75.0) | 62.9 (55.5, 69.9) | 61.6 (55.6, 67.1) |
| Fertility | 55.4 (47.1, 63.3) | 44.6 (37.5, 51.6) | 47.4 (40.2, 54.2) |
| Luteal | 68.7 (63.0, 74.2) | 87.5 (82.2, 91.9) | 73.8 (69.4, 77.9) |
| **Bleeding + position: classifier + HSMM** | | | |
| Menstrual | 80.6 (74.4, 86.6) | 65.2 (58.7, 70.7) | 68.8 (62.5, 74.5) |
| Follicular | 68.9 (62.9, 74.8) | 65.5 (58.4, 72.1) | 63.0 (57.0, 68.3) |
| Fertility | 57.2 (49.6, 65.0) | 44.4 (38.0, 50.6) | 48.6 (41.6, 55.0) |
| Luteal | 70.5 (64.8, 76.0) | 88.6 (84.2, 92.3) | 75.7 (71.8, 79.6) |
| **All four input families: classifier only** | | | |
| Menstrual | 81.4 (75.8, 86.8) | 64.2 (57.9, 69.7) | 68.8 (62.7, 74.0) |
| Follicular | 70.0 (64.3, 75.6) | 63.6 (56.7, 70.1) | 62.5 (56.9, 67.7) |
| Fertility | 54.8 (47.1, 62.5) | 44.1 (37.3, 50.8) | 47.2 (40.1, 53.8) |
| Luteal | 68.8 (63.1, 74.2) | 88.4 (84.1, 92.1) | 74.7 (70.7, 78.6) |
| **All four input families: classifier + HSMM** | | | |
| Menstrual | 81.5 (75.5, 87.4) | 65.7 (59.2, 71.3) | 69.4 (63.2, 75.0) |
| Follicular | 69.5 (63.5, 75.2) | 67.5 (60.5, 73.8) | 64.4 (58.4, 69.6) |
| Fertility | 57.0 (49.5, 64.6) | 43.9 (37.5, 50.0) | 48.4 (41.5, 54.8) |
| Luteal | 70.7 (64.9, 76.1) | 88.3 (84.1, 92.0) | 75.8 (72.0, 79.7) |

Table S3: Performance of additional classifiers. Values are participant-average macro F1 (%) with nominal 95% participant-bootstrap confidence intervals in parentheses. These comparisons use history encoding and one seed across 42 participants; the primary three-seed results are in Table 1. All four input families comprise bleeding, position, symptoms and sensors.

| Inputs | Classifier only | Classifier + HSMM |
|---|---|---|
| **Prior-only baseline** | | |
| No inputs | 12.7 (11.8, 13.6) | 15.8 (13.8, 17.9) |
| **Logistic regression** | | |
| All four input families | 63.1 (58.6, 67.4) | 63.6 (59.2, 67.8) |
| Bleeding + position + symptoms | 63.1 (58.5, 67.4) | 62.1 (57.3, 66.9) |
| Bleeding + position | 62.1 (57.5, 66.6) | 62.6 (58.2, 66.9) |
| Sensors only | 31.0 (28.0, 34.0) | 35.4 (30.3, 40.8) |
| **Random forest** | | |
| All four input families | 64.4 (60.4, 68.2) | 64.5 (60.3, 68.3) |
| Bleeding + position + symptoms | 63.2 (59.2, 67.2) | 63.2 (58.9, 67.3) |
| Bleeding + position | 64.0 (59.8, 68.0) | 63.6 (59.3, 67.6) |
| Sensors only | 26.1 (23.4, 28.8) | 32.2 (27.0, 37.8) |
| **Multilayer perceptron** | | |
| All four input families | 64.2 (60.3, 68.1) | 64.6 (60.6, 68.5) |
| Bleeding + position + symptoms | 63.4 (59.3, 67.4) | 63.9 (59.5, 68.1) |
| Bleeding + position | 63.5 (58.9, 67.8) | 64.1 (59.8, 68.2) |
| Sensors only | 27.9 (25.2, 30.7) | 29.4 (26.1, 32.5) |

Table S4: Paired baseline-model comparisons. Values are changes in macro F1 in percentage points (primary learner minus baseline), with nominal 95% participant-bootstrap intervals. Every comparison includes 42 participants and uses seed 42. Positive values favour the primary learner. Adjusted $p$ values are from two-sided sign-flip tests with Holm correction across all 16 comparisons.

| Baseline | Prediction method | Δ macro F1 (95% interval) | Adjusted $p$ |
|---|---|---|---|
| **CatBoost minus baseline** | | | |
| Prior only | Classifier only | +50.60 (+46.63, +54.20) | $< 0.001$ |
| Prior only | Classifier + HSMM | +48.70 (+44.82, +52.34) | $< 0.001$ |
| Logistic regression | Classifier only | +0.25 (-1.62, +2.13) | 1.000 |
| Logistic regression | Classifier + HSMM | +0.89 (-1.77, +3.10) | 1.000 |
| Random forest | Classifier only | -1.07 (-2.73, +0.56) | 1.000 |
| Random forest | Classifier + HSMM | +0.00 (-1.38, +1.44) | 1.000 |
| Multilayer perceptron | Classifier only | -0.88 (-2.69, +0.89) | 1.000 |
| Multilayer perceptron | Classifier + HSMM | -0.14 (-2.02, +1.72) | 1.000 |
| **Extra Trees minus baseline** | | | |
| Prior only | Classifier only | +52.14 (+48.10, +55.90) | $< 0.001$ |
| Prior only | Classifier + HSMM | +48.27 (+44.42, +51.84) | $< 0.001$ |
| Logistic regression | Classifier only | +1.79 (-0.60, +4.08) | 1.000 |
| Logistic regression | Classifier + HSMM | +0.47 (-2.15, +2.62) | 1.000 |
| Random forest | Classifier only | +0.47 (-1.39, +2.29) | 1.000 |
| Random forest | Classifier + HSMM | -0.42 (-2.11, +1.12) | 1.000 |
| Multilayer perceptron | Classifier only | +0.66 (-1.39, +2.63) | 1.000 |
| Multilayer perceptron | Classifier + HSMM | -0.57 (-2.45, +1.26) | 1.000 |

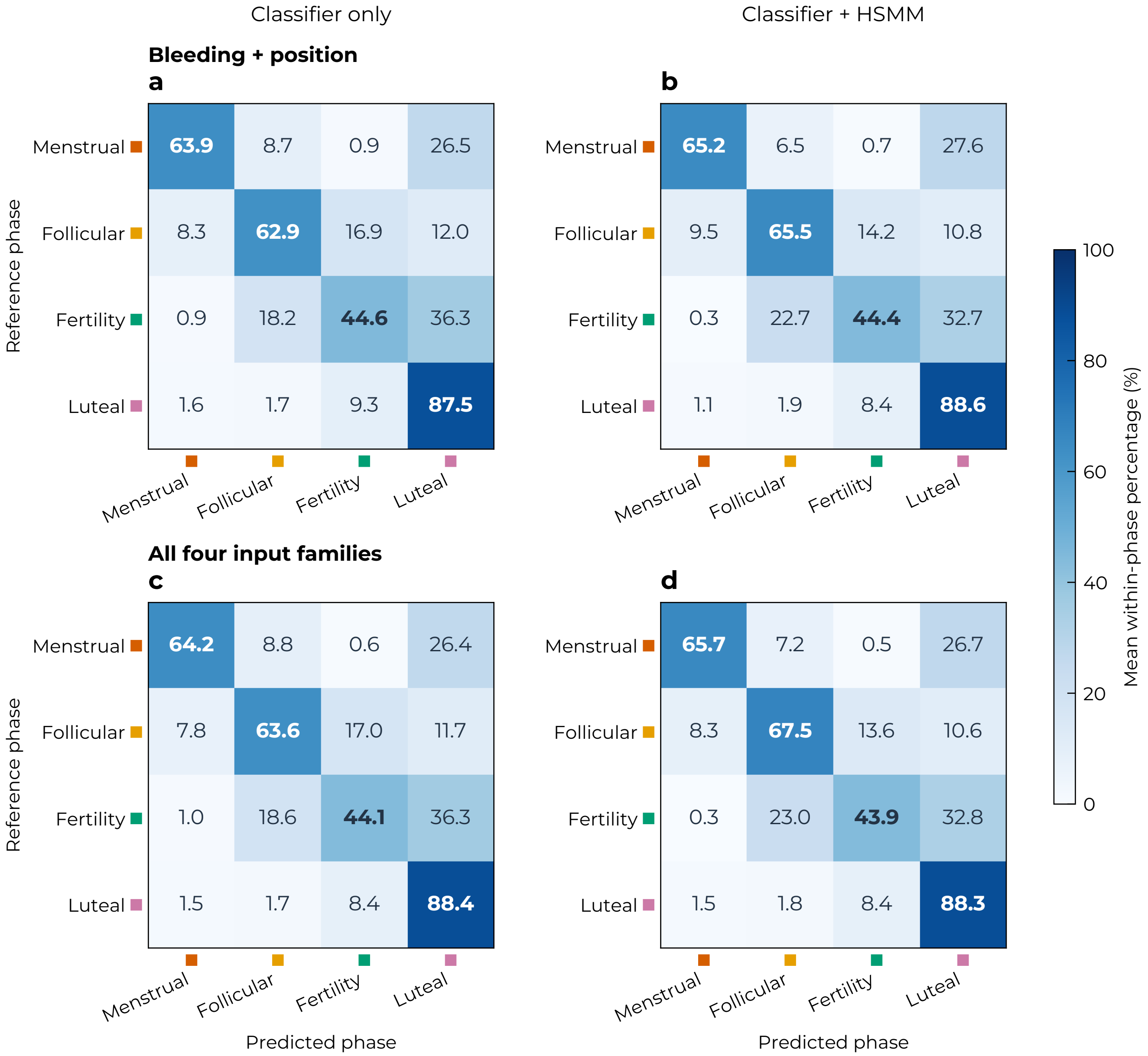


Figure S1: **Phase-classification confusion.** CatBoost with history features, using **a,b,** bleeding and position or **c,d,** all four input families. Columns compare classifier-only predictions with forward HSMM filtering. Cells show row-normalised percentages, averaged across 3 seeds within participant and then equally across participants with the reference phase (42 held-out participants overall).

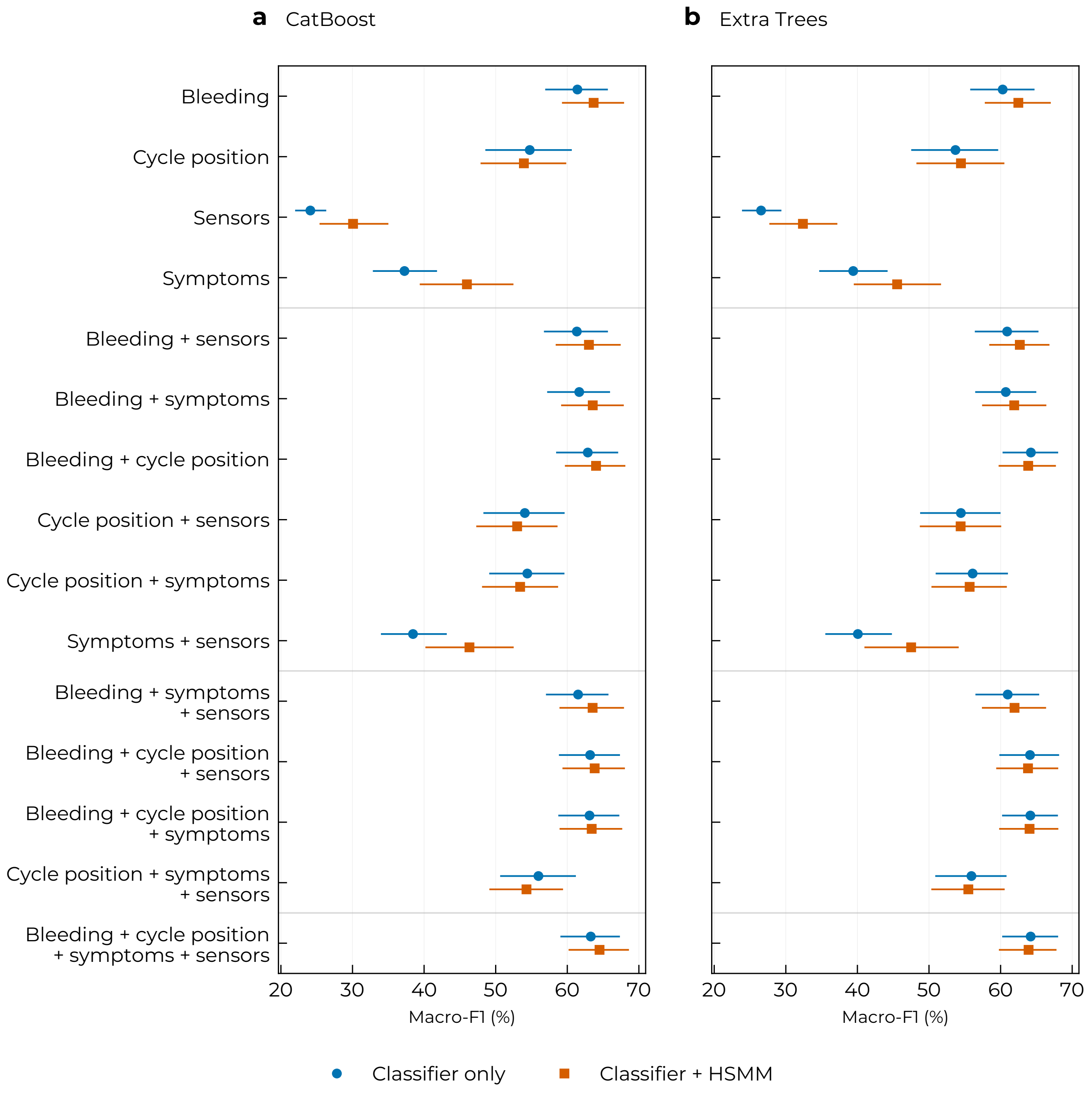


Figure S2: Performance across input combinations. Participant-average macro F1 for all 15 nonempty combinations of bleeding, cycle position, symptoms and sensors, using CatBoost (a) and Extra Trees (b) with history encoding. Circles show classifier-only predictions; squares include forward HSMM filtering. Bars indicate nominal 95% participant-bootstrap confidence intervals, with 3 seeds averaged within each of 42 participants. Rows are grouped by the number of input families.

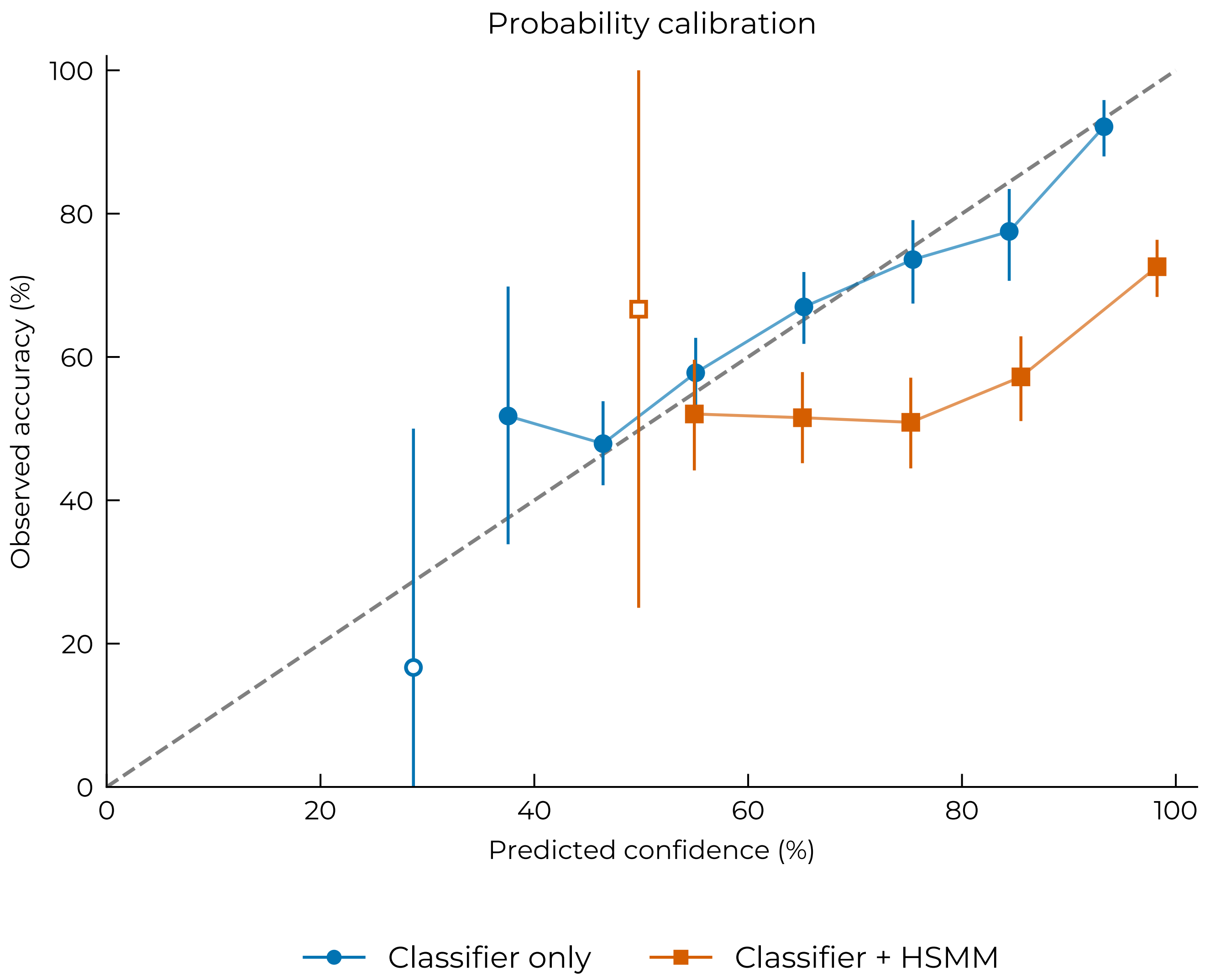


Figure S3: Calibration of phase predictions. Observed accuracy versus predicted confidence for full-input CatBoost with history encoding, before and after forward HSMM filtering. Values are averaged within each participant and confidence bin for each seed, then across available seeds within participant and across contributing participants. Bars show nominal 95% participant-bootstrap confidence intervals; the dashed line indicates ideal calibration. Unconnected open markers indicate bins with fewer than 10 participants; no interval is estimated for a bin with only one participant. All 3 seeds are included.

#### Input contributions and sensitivity analyses

Phase-specific input-family and sensor-category increments are shown in Figures S4 and S5. Figures S6 and S7 distinguish attribution within fitted models from changes after removing sensor categories and retraining. Figure S8 compares modality increments with and without nested feature selection; the procedure is described in main Methods sections "Participant-level validation and model selection" and "Input comparisons and sensitivity analyses".

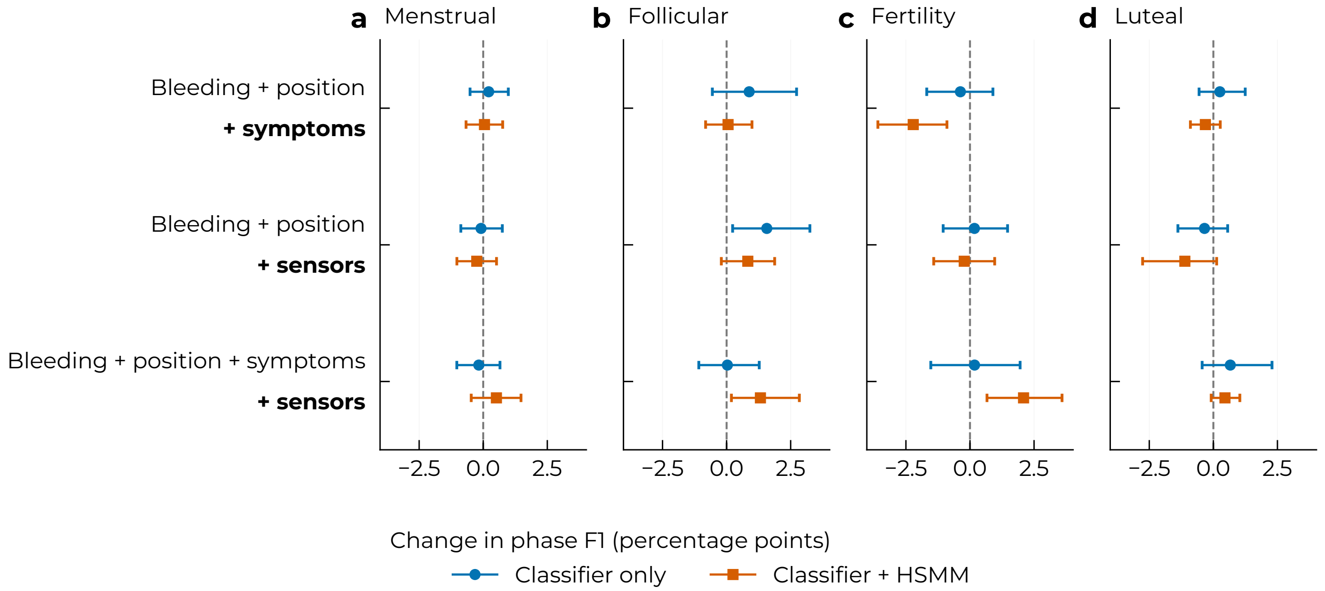


Figure S4: **Phase-specific effects of added input families. a–d,** Changes in F1 for each reference phase after adding symptoms or sensors to the input combination shown above each bold addition. Positive values favour the added inputs. All models use CatBoost with history encoding. Points show mean paired changes across 42 held-out participants after averaging 3 seeds; bars show nominal 95% participant-bootstrap intervals, without multiplicity adjustment.

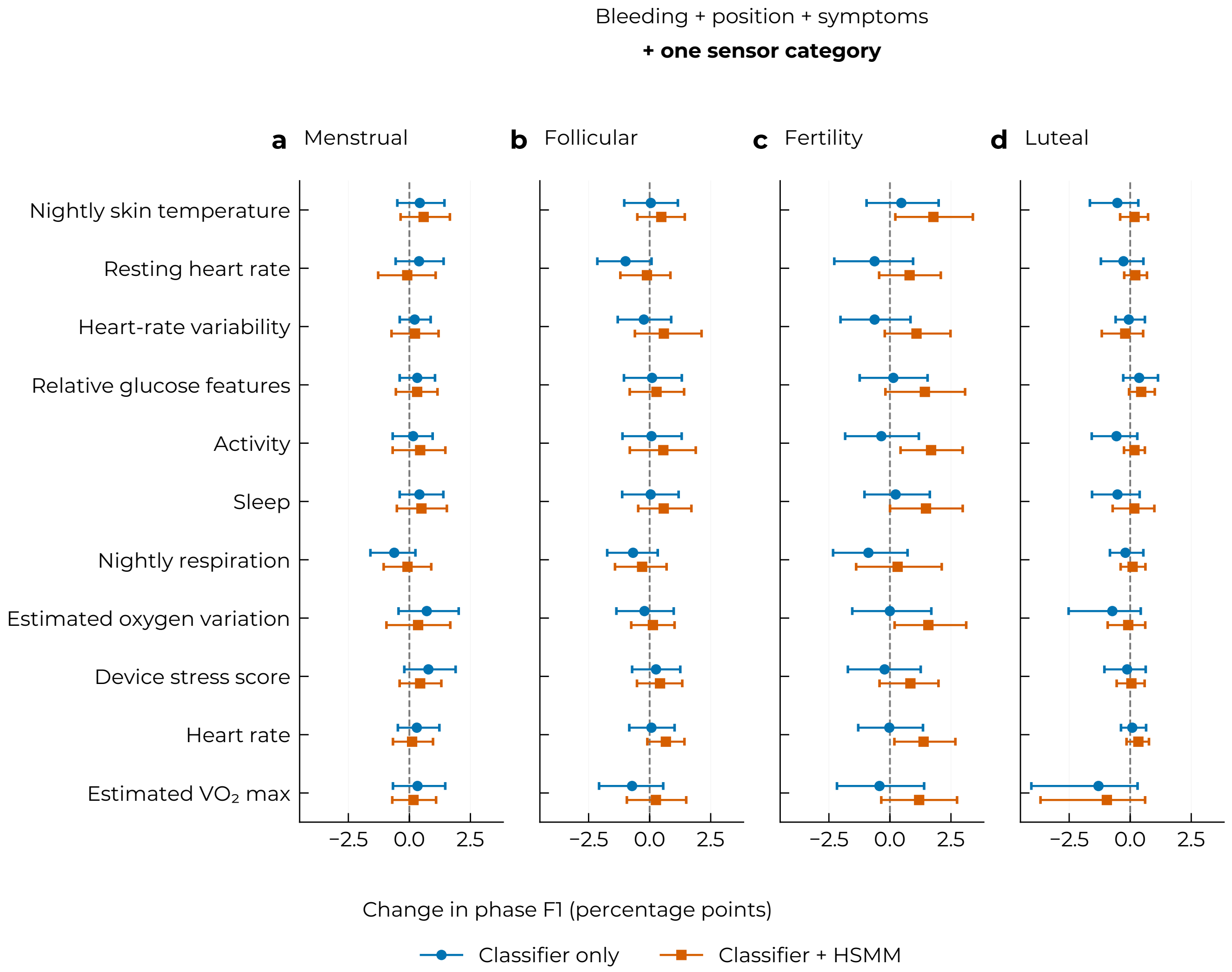


Figure S5: **Phase-specific effects of individual sensor categories. a–d,** Changes in F1 for each reference phase after adding one sensor category to bleeding, position and symptoms. Positive values favour sensor addition. Circles denote classifier-only predictions; squares include forward HSMM filtering. All models use CatBoost with history encoding. Points show mean paired changes across 42 held-out participants after averaging 3 seeds; bars show nominal 95% participant-bootstrap intervals, without multiplicity adjustment. Dashed lines mark zero.

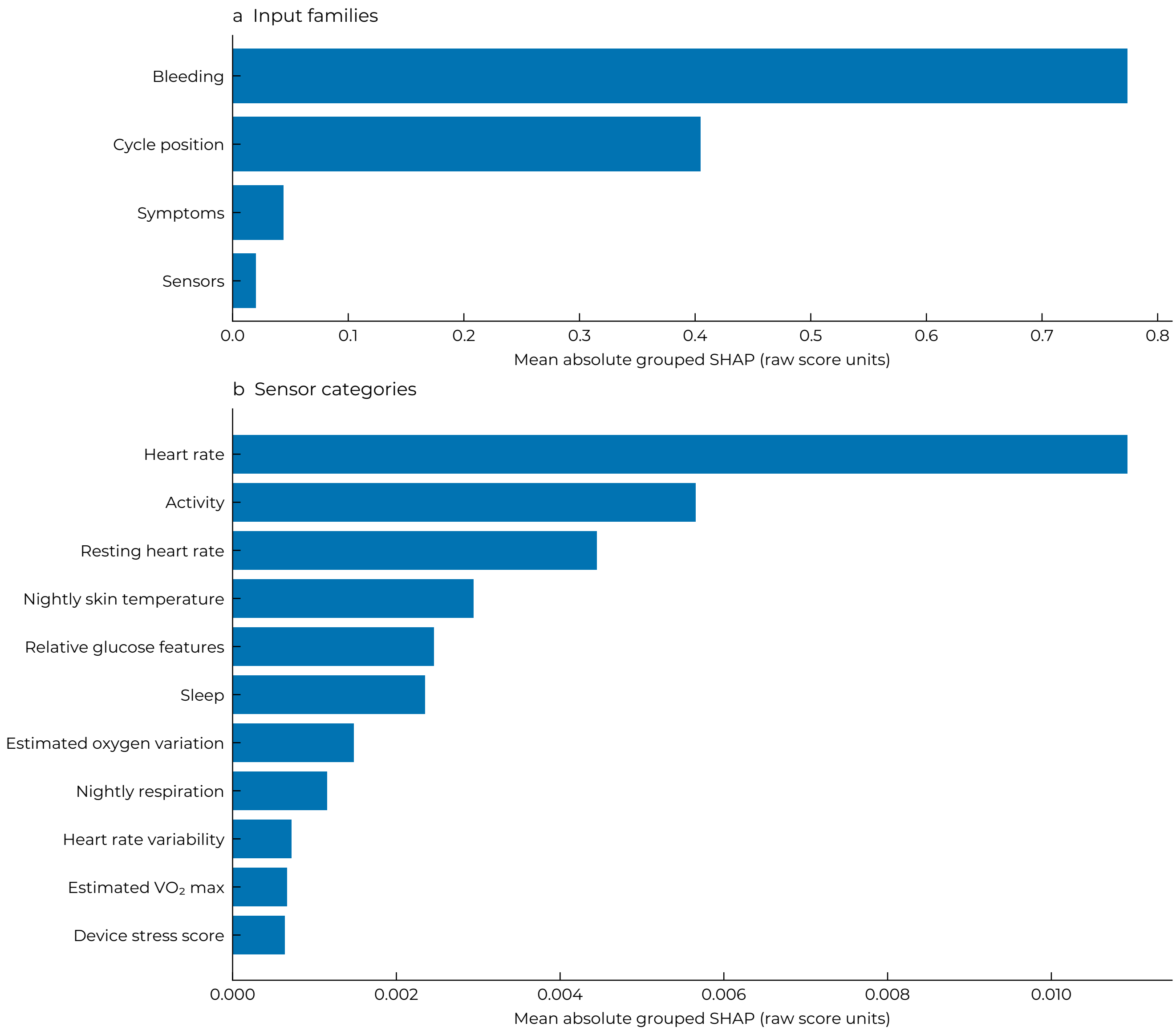


Figure S6: Grouped feature attribution. **a,** Four broad input families. **b,** Breakdown of sensor categories. Mean absolute grouped SHAP values describe contributions to raw class scores of the full-input CatBoost classifier with history encoding, before temporal filtering. Signed feature contributions are summed within each family or category before taking their magnitude, then averaged across scored days and classes, 3 seeds within participant, and equally across 42 held-out participants. Panels use the same units but different horizontal scales.

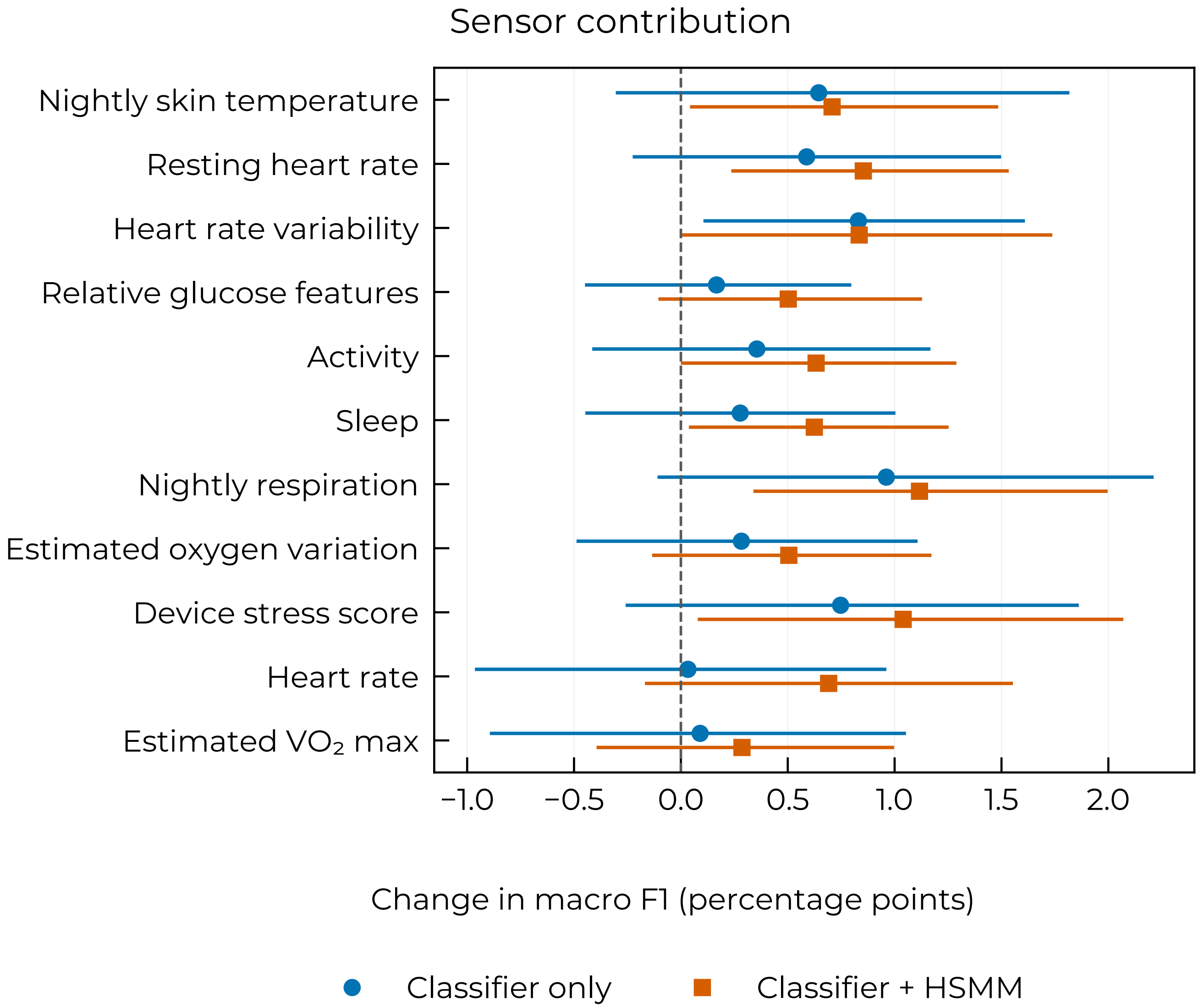


Figure S7: Conditional sensor contributions. Full-input macro F1 minus macro F1 after retraining without each sensor category, using CatBoost with history encoding. Positive values favour retaining the sensor category. Points show participant-average differences; bars show nominal 95% paired participant-bootstrap confidence intervals, with 3 seeds averaged within each of 42 participants.

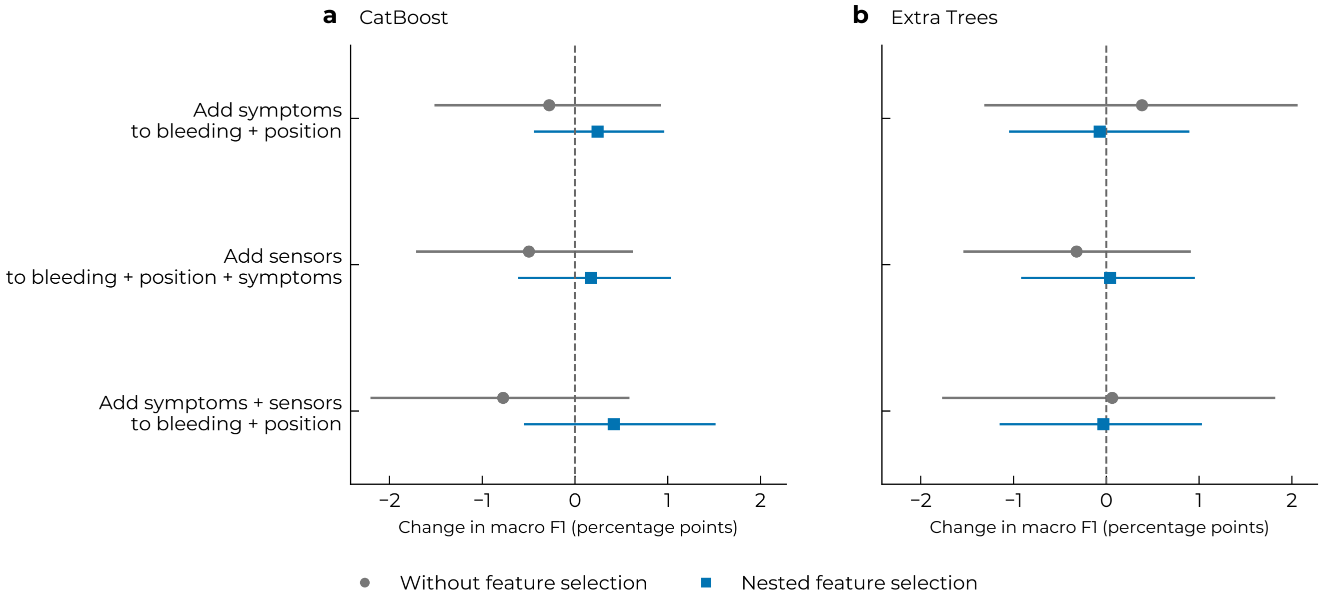


Figure S8: **Input contributions after feature selection.** **a–b,** Changes in participant-average macro F1 when adding the indicated inputs, using CatBoost and Extra Trees with history encoding. Grey circles show the matched pipeline without feature selection; blue squares show nested selection of 16, 32 or all features. Each input combination is selected and fitted independently within the same participant splits. Bars show nominal 95% paired participant-bootstrap confidence intervals across 42 participants, with 3 seeds averaged within participant. All predictions are classifier-only.

#### Temporal filtering and participant variation

Table S5 summarises participant-level performance and changes after HSMM filtering. Figures S9 and S10 show transition performance and daily probability trajectories. Table S7 reports exploratory associations with reporting completeness; the descriptors, support criteria and uncertainty calculations are defined in main Methods section "Performance measures and statistical analysis".

Table S5: Participant variation in macro F1 (%) for CatBoost with history encoding, after averaging 3 seeds within each of 42 participants. Interquartile ranges span the 25th–75th percentiles; ranges show minima and maxima. Counts compare each participant's unrounded macro F1 with and without forward HSMM filtering.

| Prediction method | Mean | Median | Interquartile range | Range |
|---|---|---|---|---|
| **Bleeding + position** | | | | |
| Classifier only | 62.9 | 64.6 | 52.5–73.5 | 33.1–82.4 |
| Classifier + HSMM | 64.0 | 67.1 | 57.0–74.7 | 29.3–84.7 |
| HSMM: 25 improved, 16 worsened, 1 unchanged | | | | |
| **All four input families** | | | | |
| Classifier only | 63.3 | 67.7 | 55.7–72.0 | 29.3–85.2 |
| Classifier + HSMM | 64.5 | 67.1 | 58.7–74.7 | 29.3–85.8 |
| HSMM: 23 improved, 17 worsened, 2 unchanged | | | | |

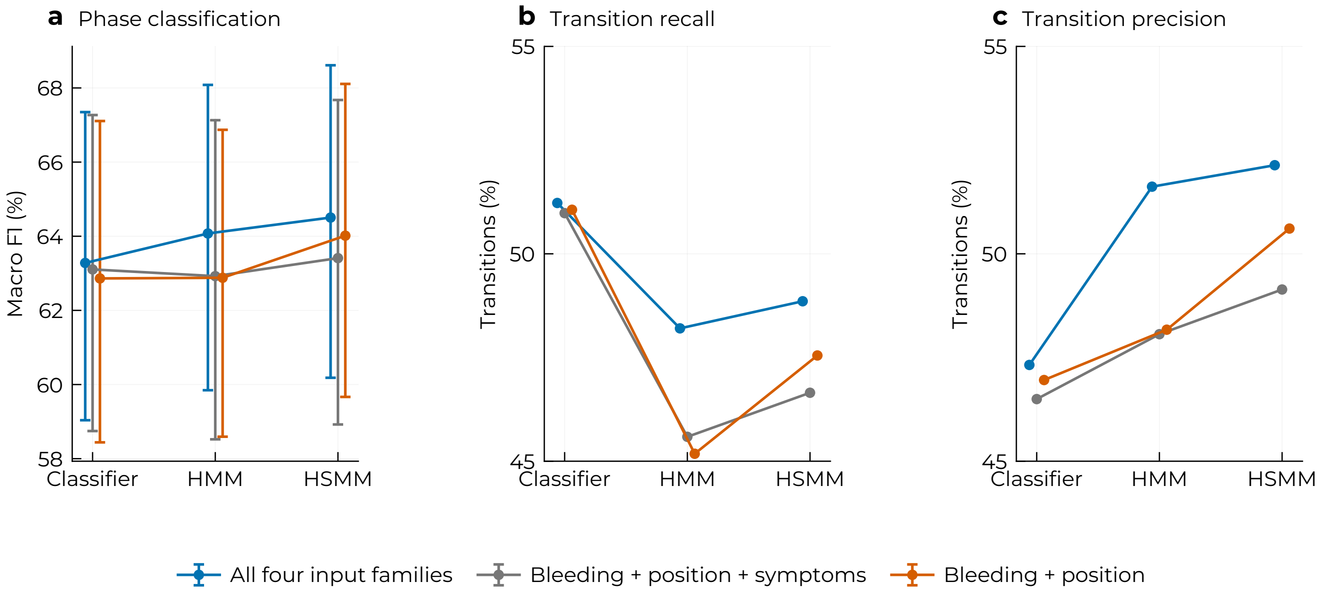


Figure S9: **Temporal filtering and phase transitions. a,** Participant-average macro F1 for CatBoost with the same fitted classifier evaluated directly or with forward HMM or HSMM filtering. Bars show nominal 95% participant-bootstrap intervals across 42 participants, after averaging 3 seeds. **b,c,** Aggregate transition recall and precision, matching transitions by destination phase within one day. Recall is the proportion of reference transitions detected; precision is the proportion of predicted transitions matched to a reference transition. Exact-day and three-day comparisons and timing errors are reported in Table S6. Transition metrics are descriptive and shown without intervals.

Table S6: Sensitivity of transition detection to matching tolerance. Cells in the exact-day and tolerance columns give precision / recall (%). MAE is the mean absolute timing error in days among matched transitions; missed transitions are excluded, and exact-day MAE is zero by definition. Predictions are unchanged across tolerances. Transitions are matched one-to-one by destination phase, excluding missing-label gaps. Counts are averaged across 3 seeds within participant and summed across 42 participants before calculating ratios and timing errors. All models use CatBoost with history encoding.

| Prediction method | Exact day | ±1 day | ±3 days | MAE (±1) | MAE (±3) |
|---|---|---|---|---|---|
| **Bleeding + position** | | | | | |
| Classifier only | 12.2 / 13.3 | 47.0 / 51.1 | 67.0 / 72.9 | 0.74 | 1.25 |
| Classifier + HMM | 13.2 / 12.4 | 48.2 / 45.2 | 75.5 / 70.8 | 0.73 | 1.32 |
| Classifier + HSMM | 13.2 / 12.4 | 50.6 / 47.5 | 77.6 / 72.9 | 0.74 | 1.32 |
| **Bleeding + position + symptoms** | | | | | |
| Classifier only | 12.7 / 13.9 | 46.5 / 51.0 | 66.2 / 72.5 | 0.73 | 1.23 |
| Classifier + HMM | 13.3 / 12.6 | 48.1 / 45.6 | 74.5 / 70.7 | 0.72 | 1.30 |
| Classifier + HSMM | 14.5 / 13.7 | 49.1 / 46.7 | 75.9 / 72.1 | 0.71 | 1.30 |
| **All four input families** | | | | | |
| Classifier only | 12.5 / 13.5 | 47.3 / 51.2 | 67.7 / 73.3 | 0.74 | 1.25 |
| Classifier + HMM | 13.4 / 12.5 | 51.6 / 48.2 | 76.4 / 71.3 | 0.74 | 1.27 |
| Classifier + HSMM | 14.0 / 13.2 | 52.1 / 48.9 | 76.9 / 72.1 | 0.73 | 1.27 |

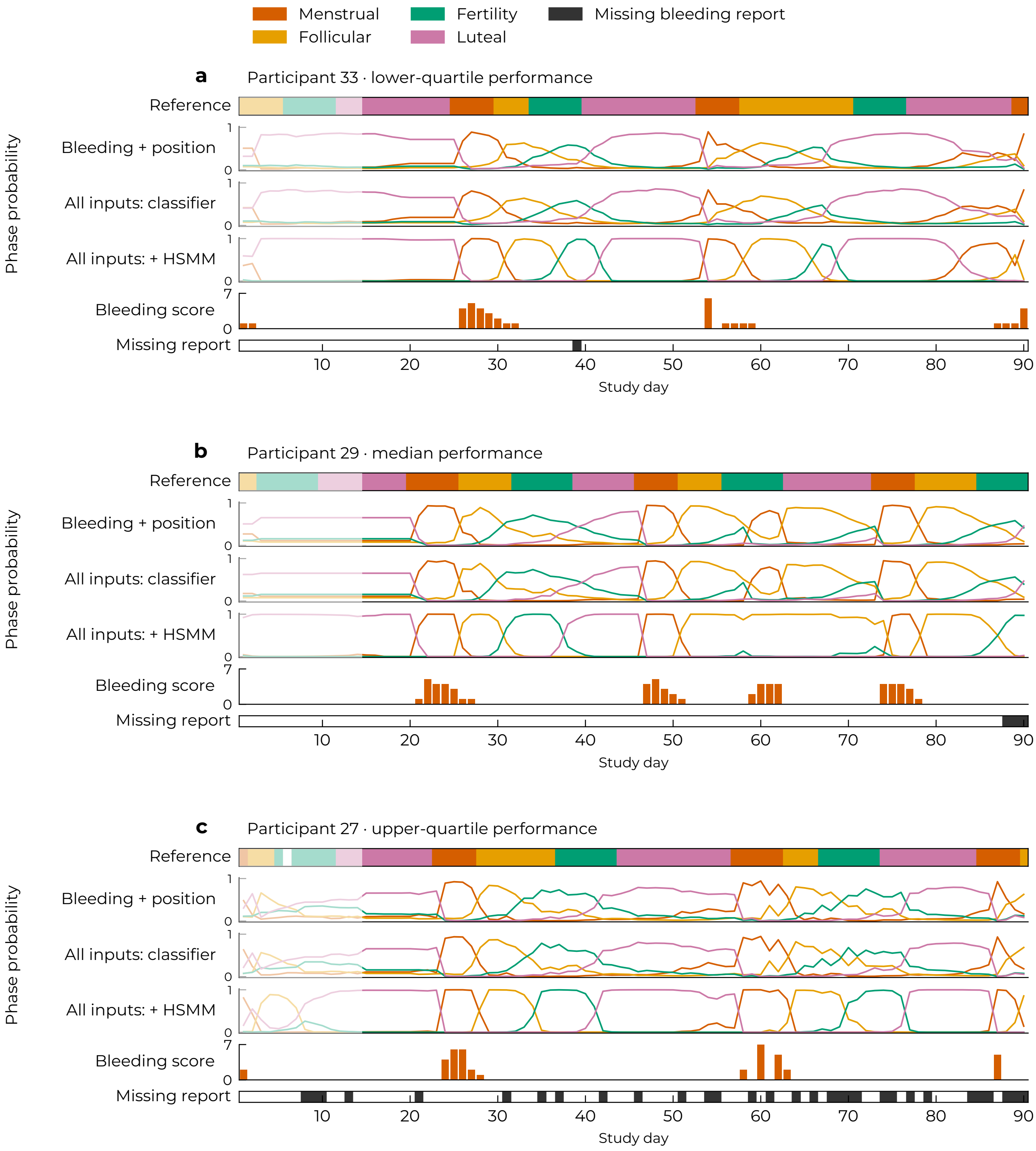


Figure S10: **Daily phase probabilities. a–c,** Participants nearest the 25th, 50th and 75th percentiles of macro F1 for full-input CatBoost with HSMM filtering, averaged across 3 seeds. Curves show held-out phase probabilities (0–1) from the bleeding-and-position classifier, full-input classifier and full-input classifier with forward HSMM filtering. Full inputs comprise bleeding, position, symptoms and sensors; models use history encoding and seed 42. Pale regions mark unscored days. Bars show bleeding intensity (0–7); dark cells identify missing bleeding reports.

Table S7: Relationships between participant characteristics and prediction performance. Values are Spearman correlations with nominal 95% participant-bootstrap confidence intervals. All rows include 42 participants; scores are averaged across 3 seeds within participant. Positive correlations indicate higher performance in A or a more favourable change in B–C as the characteristic increases; negative correlations indicate the reverse. These are associations across participants, not changes in F1 themselves. Intervals spanning zero leave the direction uncertain. Correlations with scored-day count were not estimated because counts varied too little; cycle-length variability had sufficient observations for only three participants. Intervals are exploratory and are not adjusted for multiple comparisons.

| Participant characteristic | CatBoost | Extra Trees |
|---|---|---|
| **A. Which participants have higher classification performance?** | | |
| Outcome: full-input classifier macro F1. | | |
| More missing bleeding reports | -0.21 (-0.49, +0.11) | -0.25 (-0.55, +0.09) |
| More missing symptom reports | -0.15 (-0.44, +0.17) | -0.18 (-0.47, +0.15) |
| Longer gaps in bleeding reports | -0.08 (-0.39, +0.23) | -0.13 (-0.45, +0.20) |
| More complete reference cycles | +0.12 (-0.17, +0.39) | +0.29 (+0.02, +0.52) |
| **B. Do sensors help more when reports are missing?** | | |
| Outcome: change in macro F1 after adding sensors to bleeding, position and symptoms. | | |
| More missing bleeding reports | -0.11 (-0.43, +0.21) | -0.16 (-0.46, +0.16) |
| More missing symptom reports | -0.06 (-0.38, +0.25) | -0.22 (-0.50, +0.10) |
| **C. Does HSMM filtering help more when reports are missing?** | | |
| Outcome: change in macro F1 after applying HSMM to the full-input classifier. | | |
| More missing bleeding reports | +0.26 (-0.07, +0.57) | +0.45 (+0.15, +0.69) |
| More missing symptom reports | +0.17 (-0.15, +0.47) | +0.27 (-0.05, +0.55) |

### Cohort summaries and feature dictionary

Tables S8 and S9 summarise observation coverage, annotation-defined durations and reporting completeness. Table S10 lists the input measurements and their source tables and units in mcPHASES [1]. Definitions and processing criteria are given in main Methods sections "Dataset and reference phases" and "Input families and feature construction". The accompanying code exports the full engineered-feature inventory and fold-specific selections.

Table S8: Observation coverage and reference-phase durations. IQR denotes the interquartile range. The count $n$ represents participants for per-participant summaries, cycles for cycle lengths, and segments for phase durations. Complete phase segments have observed neighbouring labels at both boundaries; censored durations are observed portions only. Complete cycles span two observed menstrual onsets with the ordered Menstrual, Follicular, Fertility and Luteal sequence and no missing labels. Boundary fragments are not counted as complete cycles.

| Measure | $n$ | Median (IQR) | Range |
|---|---|---|---|
| Observation days per participant | 42 | 90 (90–90) | 38–90 |
| Scored days per participant | 42 | 76 (76–76) | 24–76 |
| Days with any report per participant | 42 | 89 (86–90) | 19–90 |
| Complete cycles per participant | 42 | 2 (1.25–2) | 0–4 |
| Complete reference-cycle length (days) | 76 | 29 (27–33) | 11–42 |
| **Phase durations (days)** | | | |
| Menstrual (complete) | 113 | 6 (4–7) | 1–15 |
| Menstrual (censored) | 16 | 4 (2–6.25) | 1–7 |
| Follicular (complete) | 111 | 7 (4–10) | 2–23 |
| Follicular (censored) | 26 | 5 (3–7) | 1–14 |
| Fertility (complete) | 114 | 7 (6–7) | 3–7 |
| Fertility (censored) | 19 | 4 (2–5) | 1–7 |
| Luteal (complete) | 108 | 10.5 (9–12) | 1–15 |
| Luteal (censored) | 25 | 6 (3–9) | 1–37 |
| **Reference-sequence coverage** | | | |
| Complete cycles | 76 | – | – |
| Observation-boundary fragments | 84 | – | – |
| Bounded intervals with missing labels | 0 | – | – |
| Bounded intervals with noncanonical order | 0 | – | – |

An observed onset requires a non-Menstrual label on the preceding day. We retained 2 annotation-defined cycles lasting 11 and 13 days. The reference series contains 1 missing label; adjacent phase segments are censored. The 84 observation-boundary fragments represent partial labelled records rather than confirmed physiological cycles.

Table S9: Missing reports and consecutive reporting gaps. **A,** Missing participant-days, n (%), across the full observation period (3,698 participant-days) and the evaluation period after warmup (3,110 participant-days). Zero and "Not at all" responses are treated as observed. **B,** Consecutive reporting gaps, summarised by the number of affected participants, total number of gaps, counts by duration and longest observed gap in days. Gaps are counted separately within participants. Gap definitions are not mutually exclusive.

**A. Missing reports on individual days**

| Report variable | All observation days Missing, n (%) | Evaluated days Missing, n (%) |
|---|---|---|
| Bleeding volume | 509 (13.8) | 475 (15.3) |
| Bleeding colour | 509 (13.8) | 475 (15.3) |
| Appetite | 378 (10.2) | 353 (11.4) |
| Exercise level | 378 (10.2) | 353 (11.4) |
| Headaches | 378 (10.2) | 353 (11.4) |
| Cramps | 378 (10.2) | 353 (11.4) |
| Sore breasts | 378 (10.2) | 353 (11.4) |
| Fatigue | 378 (10.2) | 353 (11.4) |
| Sleep issues | 378 (10.2) | 353 (11.4) |
| Mood swings | 378 (10.2) | 353 (11.4) |
| Stress | 378 (10.2) | 353 (11.4) |
| Food cravings | 378 (10.2) | 353 (11.4) |
| Indigestion | 378 (10.2) | 353 (11.4) |
| Bloating | 378 (10.2) | 353 (11.4) |
| All 14 reports missing | 152 (4.1) | 145 (4.7) |
| At least one report missing | 735 (19.9) | 683 (22.0) |

**B. Separate gaps of consecutive missing days**

| Gap definition | Participants, n | Separate gaps, n | Gaps lasting 1–4 days, n | Gaps lasting ≥5 days, n | Longest gap, days |
|---|---|---|---|---|---|
| All 14 reports missing | 23 | 60 | 55 | 5 | 20 |
| Bleeding volume missing | 38 | 333 | 325 | 8 | 20 |
| At least one report missing | 41 | 301 | 273 | 28 | 23 |

Gaps touching the start or end of recording are included; their observed lengths may underestimate the full gap. 16 all-report gaps, 17 bleeding-volume gaps and 38 any-report gaps touch a boundary. Cycle completeness is determined by reference-label availability, independently of reporting gaps.

Table S10: Feature dictionary. Counts in parentheses sum to 69 daily base features: 4 position, 10 bleeding, 12 symptom and 43 sensor features. Source names identify the mcPHASES tables. Availability indicators and temporal expansions are excluded from these counts. Processing criteria are specified in main Methods section "Input families and feature construction".

| Input and source | Measurements and units |
|---|---|
| Position (4)<br>Bleeding-derived | Days since detected bleeding onset; sine and cosine of position in a 28-day reference cycle; clipped elapsed-cycle position. Days and dimensionless encodings. |
| Bleeding (10)<br>`hormones_and_selfreport` | Flow volume on an ordinal 0–7 scale (not at all to very heavy); nine nominal flow-colour categories encoded as binary 0/1 indicators. |
| Symptoms (12)<br>`hormones_and_selfreport` | Appetite, exercise level, headaches, cramps, sore breasts, fatigue, sleep issues, mood swings, stress, food cravings, indigestion and bloating. Responses were encoded on a common ordinal scale from 0 (not at all) to 5 (very high). |
| Skin temperature (1)<br>`computed_temperature` | Nightly skin temperature in $^{\circ}$C. |
| Resting heart rate (1)<br>`resting_heart_rate` | Device-reported resting heart rate in beats/min. |
| Heart-rate variability (1)<br>`heart_rate_variability_details` | Daily median root mean square of successive differences (RMSSD), measured in ms; entered as $\ln[1 + \mathrm{RMSSD}/(1\,\mathrm{ms})]$ (dimensionless). |
| Glucose (5)<br>`glucose` | SD/mean; interquartile range/median; 90th/10th percentile ratio; daily mean divided by the preceding seven-day mean; and observed fraction. All five measures are dimensionless. |
| Activity (16)<br>`active_minutes`, `time_in_heart_rate_zones`, `steps`, `distance`, `calories`, `altitude` | Four activity-intensity durations and four heart-rate-zone durations in minutes; recorded steps, distance in metres, calories in source units and altitude gain in metres, each with an observed-minute count (minutes). |
| Sleep (5)<br>`sleep`, `sleep_score` | Minutes asleep, minutes awake, time in bed, efficiency (%) and device sleep score (points). |
| Respiration (2)<br>`respiratory_rate_summary` | Full-night breathing rate and its SD, in breaths/min. |
| Oxygen variation (5)<br>`estimated_oxygen_variation` | Mean, SD, median and interquartile range of the exported signed signal, plus observed-minute count. Signal values are not $SpO_2$ percentages. |
| Device stress (1)<br>`stress_score` | Device-generated stress-management score (points). Distinct from self-reported stress. |
| Heart rate (5)<br>`heart_rate` | Mean, SD, median and interquartile range in beats/min, plus observed-minute count. |
| Estimated fitness (1)<br>`demographic_vo2_max` | Device-estimated $VO_2$ max in $\mathrm{ml\,kg^{-1}\,min^{-1}}$. |

Additional predictors, excluded from the base-feature counts, comprise binary availability indicators (0/1); 7- and 14-day rolling means, sample SDs and observed fractions for non-position inputs; and, in the dynamics representation, 1-, 3- and 7-day differences, a 7-day slope and deviation from the expanding prior mean. Means, SDs, differences and deviations retain the base predictor's units; slopes are expressed per day. Availability indicators and observed fractions are dimensionless. Construction and fold-specific selection are described in main Methods sections "Input families and feature construction" and "Participant-level validation and model selection".
